# RAGE mediates LPS-induced inflammatory pain in human skin

**DOI:** 10.64898/2025.12.18.25342498

**Authors:** Felix J. Resch, Sibylle Pramhas, Bernd Jilma, Sabine Sator, Stefan Heber, Michael J.M. Fischer

## Abstract

Inflammatory pain is a major clinical challenge, yet appropriate human models mimicking local infection are lacking. We established a novel human pain model based on intradermal administration of lipopolysaccharide (LPS), a component of Gram-negative bacteria, and investigated the time course and underlying molecular mechanisms of inflammatory pain hypersensitivity. In a placebo-controlled pilot study with 12 healthy subjects, the intradermal LPS injection induced hyperaemia peaking at 4.5 h and mechanical hypersensitivity peaking at 6 h. Hypersensitivity to increasingly acidic injections and mechanical pinch lasted longer than hyperaemia. The double-blind, randomized, placebo-controlled full crossover main study was completed by 40 subjects and investigated the role of the Receptor for Advanced Glycation End-products (RAGE). Co-injection of the RAGE antagonist azeliragon largely reduced LPS-induced hyperaemia (-87%) and significantly attenuated hypersensitivity to mechanical (-55%) and increasingly acidic stimuli (-40%). In contrast, the Toll-like receptor 4 antagonist resatorvid had no effect on any readout. In both naïve and inflamed skin, TRPV1 antagonist BCTC inhibited the majority of acid-induced pain. LPS-induced inflammation caused a substantial shift in the pH sensitivity of pain, suggesting that even mild tissue acidification contributes to inflammatory pain. The human LPS skin inflammation model is largely RAGE-dependent, highlighting its potential as a target in inflammation.

## Introduction

Given that most types of pain have an inflammatory component and most pain conditions involve inflammation, robust human models are essential for elucidating the underlying mechanisms. There are several sterile skin inflammation models, including UV-B radiation^1^, the proinflammatory mediators NGF^2^ or bradykinin^3^. Further, freeze injury^4^ or burn injury models^5^ have been developed, which also have an inflammatory component. However, there are currently no human pain models mimicking local infection. Intravenous administration of Lipopolysaccharide (LPS), a highly pro-inflammatory component of gram-negative bacteria, is an established human model of systemic inflammation^6^. LPS has also been applied intradermally, resulting in small-diameter inflamed skin lesions^7^, which appears suitable for establishing a pain model.

In cellular as well as in animal models, LPS has been shown to sensitize, and is generally thought to be mediated by Toll-like receptor 4 (TLR4)^8^, a discovery honoured by the 2011 Nobel prize in physiology or medicine^9^. More recently, a potential role of receptor for advanced glycation end-products (RAGE) pathway has been suggested^10,11^. This study makes use of the RAGE antagonist azeliragon which has reached phase III (clinical trials NCT02080364, NCT02916056 and NCT05815485) and the TRL4 antagonist resatorvid to clarify the relative importance of these receptors in LPS-induced human skin inflammation.

Inflammation is associated with tissue acidification^12^. Sensory neurons express several ion channels, which have been shown to be sensitized or activated by acidic pH, including transient receptor potential cation channel subfamily V member 1 (TRPV1)^13^ and subfamily A 1 (TRPA1)^14^, or acid-sensing ion channels (ASICs)^15^. In healthy human skin, TRPV1 is responsible for the majority of acid-induced pain^16^. Inflammatory mediators, like ATP^17^, PGE ^18^, bradykinin^19^ or NGF^20^ sensitize proton activation of TRPV1 *in vitro*, which suggests a relevant role of this ion channel in inflammation. Acid-induced pain has not been investigated in inflammation. It is unclear whether TRPV1 continues to be the primary acid sensor in inflamed human skin.

The objectives of this study were 1) to establish LPS-based skin inflammation as a novel human pain model, 2) to elucidate the underlying pathway leading to inflammatory pain hypersensitivity and 3) to clarify the role of TRPV1 in acid-induced pain in LPS-based skin inflammation. To address objective 1, a pilot study involving 12 healthy subjects was conducted to measure the time course of LPS-induced hyperaemia and pain hypersensitivity. Mechanical pinch, contact heat and intradermal injection of acidic fluid were used as noxious stimuli. Based on these data, 40 healthy subjects completed the main study to address objectives 2 and 3 by pharmacological means.

## Results

### Time course of intradermal LPS-induced inflammation

The time course of hyperaemia and pain hypersensitivity, triggered by intradermal LPS application, was assessed in 6 female and 6 male volunteers with a median age of 23 years and a range of 20 to 29 years (Fig. 1A). Both hyperaemia, measured by laser speckle imaging, and hypersensitivity to increasingly acidic intradermal injection, rated on a 0-100 numerical scale, peaked at 4.5 h after LPS injection (Fig. 1B,C,F). Hypersensitivity to mechanical pinch had its peak at 6 h after LPS injection, but was close to the maximum after 4.5 h (Fig. 1E). Thus, the 4.5 h time point of the pilot study appeared most suitable for the main study. The heat pain threshold was surprisingly not altered to a relevant extent by inflammation (Fig. 1D) and was therefore omitted for the main study. The increased sensitivity to increasingly acidic injections due to LPS-induced inflammation receded more slowly than other modalities (Fig. 1F,S1). After 50 h the hyperaemia had subsided. However, compared to the peak sensitivity, after 50 h about half the mechanical hypersensitivity remained and the majority of the hypersensitivity to acid.

**Figure 1:**
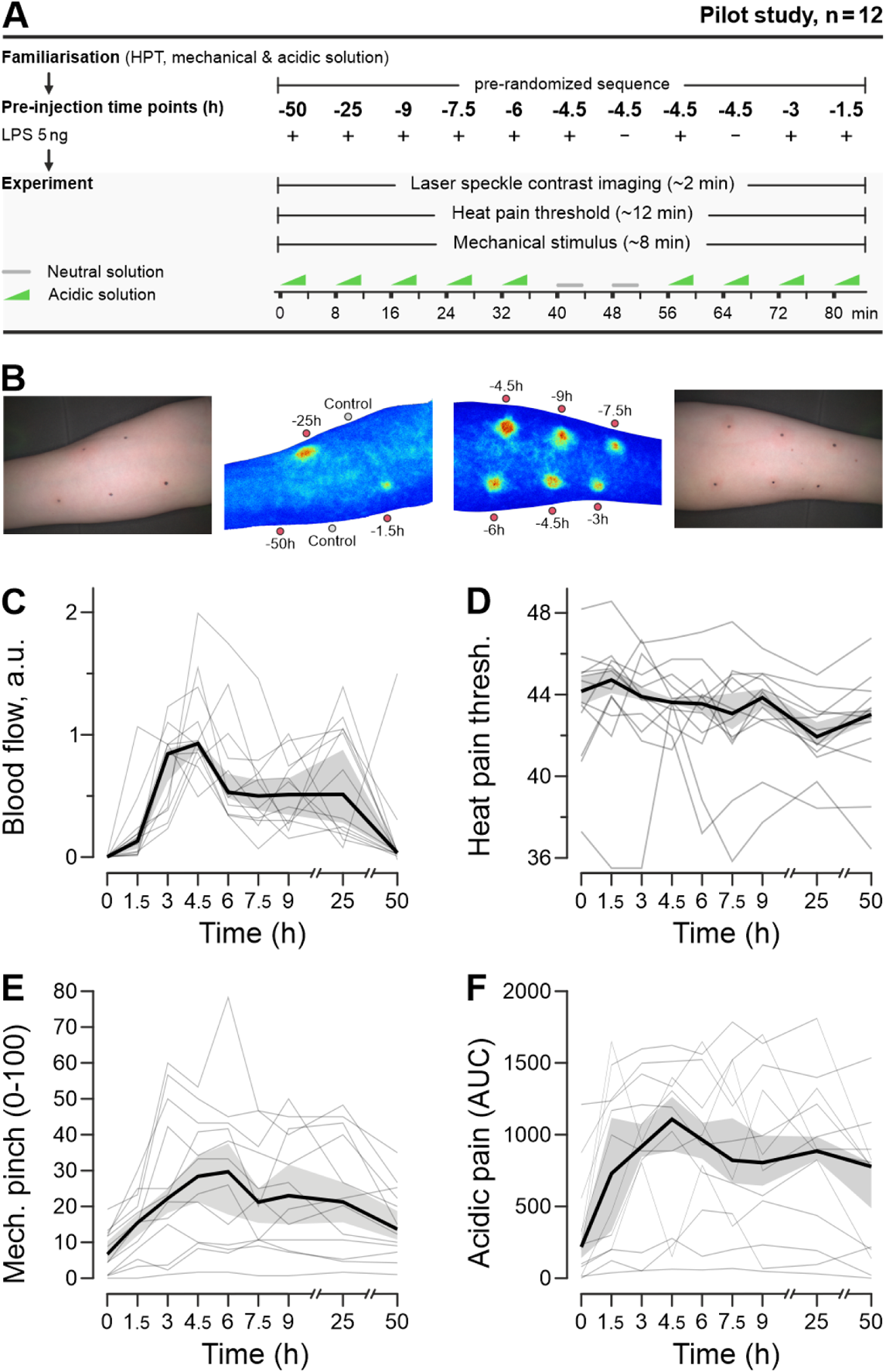
Time course of hyperaemia and pain hypersensitivity in the LPS inflammation model. **A)** Protocol of the pilot study, all 12 participants completed the protocol. **B)** Image and laser speckle-based blood flow imaging of a representative subject. LPS was injected 1.5-50 h earlier as indicated. Blood flow was acquired before subsequent testing. Due to injections with and without acid thereafter, the blood flow without LPS treatment and the blood flow 4.5 h after LPS was measured twice in each of the 12 subjects. **C)** Hyperaemia is detectable at 1.5 h, peaks at 4.5 h after LPS injection, and recovers within 50 h to baseline. **D)** Heat pain threshold (in °C) was not systematically altered by LPS-induced inflammation. **E)** Numerical rating of the pain intensity induced by a 2560 mN mechanical pinch stimulus application reaches a peak at 6 h. **F)** AUC pain induced by increasingly acidic injections. In panels C-F, individual traces of all 12 participants are shown by grey lines, the median in black, and 68% CI is shaded in grey. Data for the two spots without LPS (i.e. time point = 0) and the two spots 4.5 h after LPS application were averaged.

**Figure S1.**
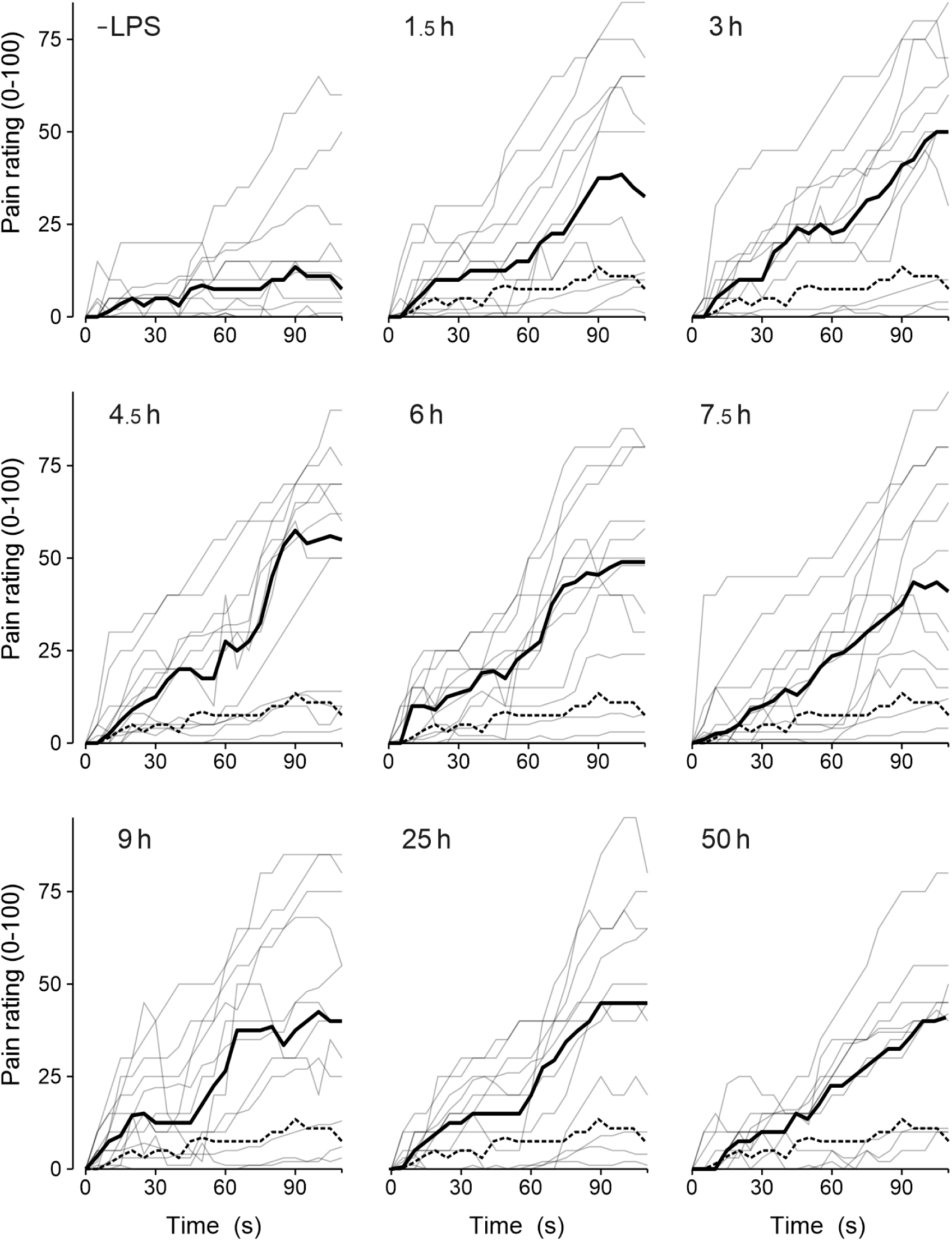
Time course of LPS-induced acid sensitisation. For all 12 participants of the pilot study, the pain due to the increasingly acidic injection is visualized. Compared to the observations without LPS (-LPS), the period after intradermal injection of 5 ng LPS is provided. All panels visualize the median (black line) and the median obtained without LPS provide a reference (dotted line). Note that there is a gradual increase until 4.5 h after LPS, and a rather stable level in the period 4.5–9 h after LPS.

### LPS-induced hyperaemia depends on RAGE, but not TLR4

The main study had 42 participants, 20 female and 20 male subjects with a median age of 26 years and a range of 20-40 years completed the protocol (Fig. 2A). In two participants the protocol was not completed (data not further considered), when it became apparent that the order of the pre-specified sequence was not followed correctly. The blood flow 4.5 h after control injection was 0.008 arbitrary units (CI 0.005–0.011), which is similar to non-injected skin. LPS increased the blood flow after 4.5 h to 1.26 (CI 1.12–1.40, Fig. 2B,C), which is an increase similar to the pilot study. The primary endpoint of the main study was the comparison of hyperaemia in skin spots injected with LPS alone vs. RAGE antagonist azeliragon coinjected with LPS. Compared to LPS alone, the addition of azeliragon reduced skin hyperaemia by 87% to 0.16 (CI 0.13–0.20, p < 0.0001, Fig. 2D). In contrast, the TLR4 antagonist resatorvid did not alter skin hyperaemia (105% of LPS alone, mean estimate 1.32, CI 1.15–1.50, Fig. 2E). Exploratory addition of sex to the mixed model did not indicate sex-dependent differences in blood flow (‘sex’*’injection type’ p = 0.44).

**Figure 2:**
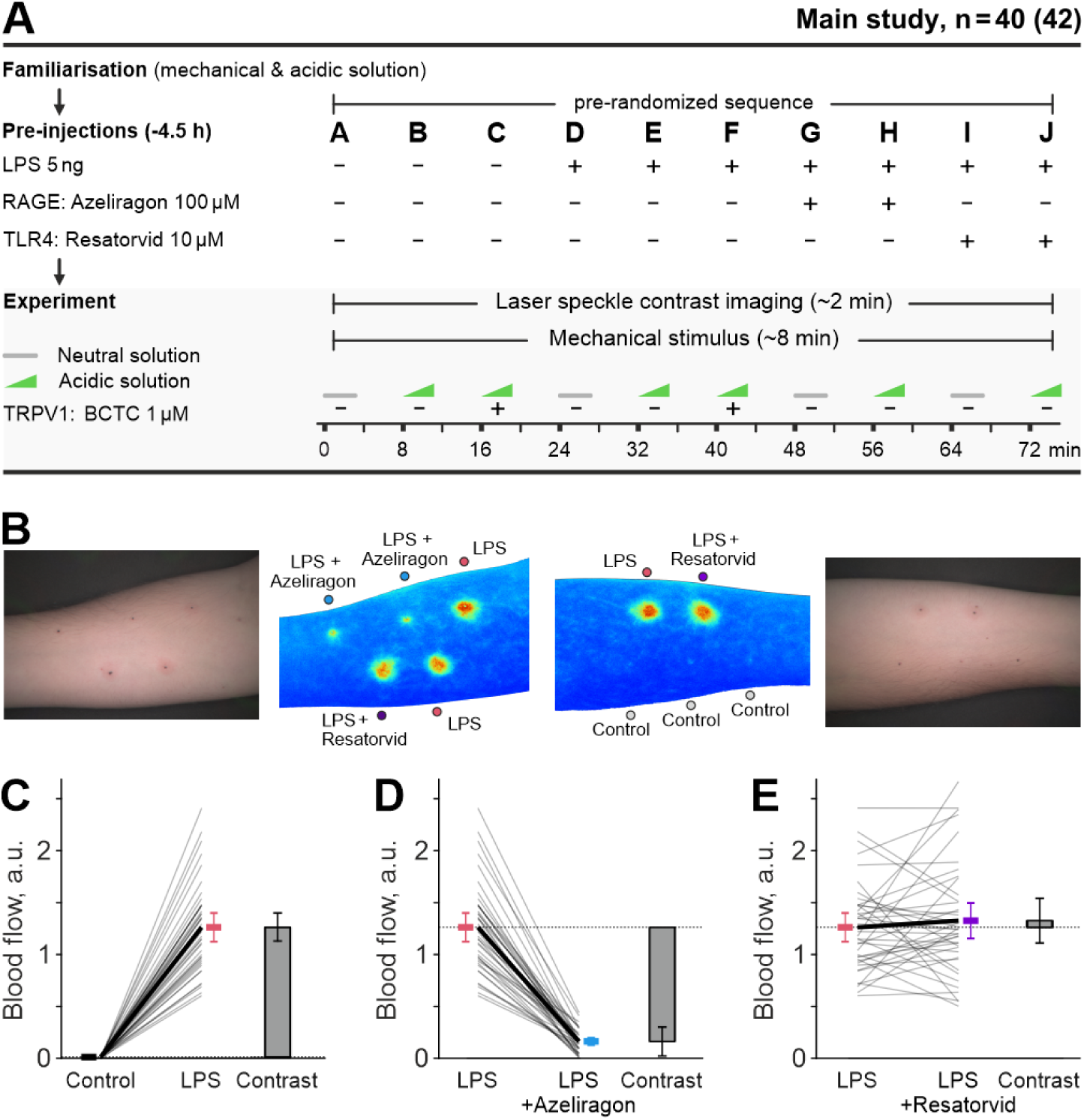
Hyperaemia in the LPS inflammation model largely depends on RAGE, but not TLR4. **A)** Protocol of the main study with 42 participants, 40 completed the protocol. **B)** Representative subject for laser speckle-based blood flow imaging at 4.5 h after control injections without LPS and LPS injections, both measured in three spots. RAGE antagonist azeliragon and TLR4 antagonist resatorvid were coapplied with LPS in two spots each. **C)** Blood flow is increased in all LPS-injected spots in all 40 subjects. Mean estimates are provided with 95% CIs and the contrast between the two groups. **D)** In comparison to LPS-induced hyperaemia, coapplication of azeliragon 100 µM reduced blood flow by 87% (CI 84–89%), which is the primary endpoint of the main study. **E)** In contrast, the mean and confidence interval after coapplication of resatorvid 10 µM indicated no relevant dependence of blood flow on TLR4.

### LPS-induced inflammation sensitizes acid-induced pain

The acid-induced pain model was refined from the previous models, which used a constant pH21 or three acidic levels16, to a pH ramp with a largely linear decrease from a pH of 7.24 to 5.44 in the current model (Fig. 3A-C). Pairwise comparison shows that in naïve skin increasingly acidic injections were more painful than pH 7.4 injections (Fig. 3D-F), pH 7.4 injections were more painful in inflamed skin compared to naïve skin (Fig. 3D,G,J), and increasingly acidic injections were more painful in inflamed skin than in naïve skin (Fig. 3F,I,L), and also compared to pH 7.4 injections in inflamed skin (Fig. 3J,K,L). Addressing the exploratory hypothesis, the difference in pain between increasingly acidic and pH 7.4 injections was indeed greater in inflamed vs. naïve skin (interaction ‘acid’*’LPS’ p < 0.001).

**Figure 3:**
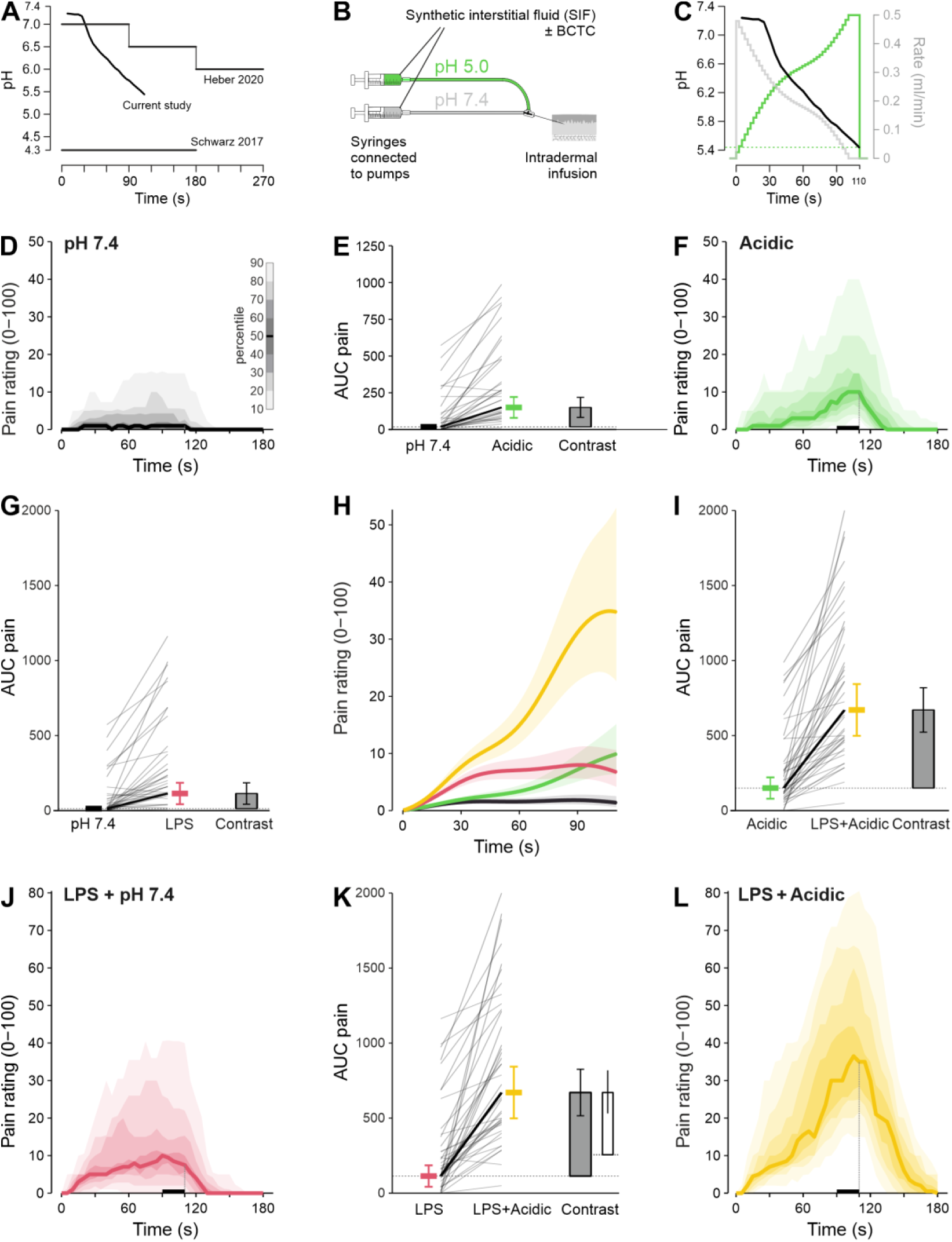
LPS-induced inflammation increases acid-induced pain. **A)** Acid stimulation in prior studies, from a fixed acidic pH 4.3, to three acidic levels, to the pH ramp of the current study. **B)** Improved acid-induced pain model with increasingly acidic pH, mixed from a pH 7.4 and 5.0 fluid. **C)** Nonlinear adaptation of the rates of two pumps results in a largely linear pH ramp (mean of three measurements). The time is aligned with the panel below. **D,F,J,L)** Time courses of pain ratings, the distribution is visualized with the median as a solid line and decreasing grey or colour shades for percentiles more distant from the median in 10% percentile steps, as indicated by the scale bar in panel D. The injection lasts 110 s, the end is visualized by a vertical dashed line. AUC pain is calculated for the period 90–110 s, marked by a bar on the x-axis. **E,G,I,K).** Mean estimates for the AUC pain, spaghetti plot and contrast between the two groups, for which the time course is given in the two adjacent diagrams. Panel K also visualizes the contrast (open bar) for the prespecified hypothesis that pain induced by acid in addition to mechanically induced pain is greater after LPS injection than after control injection. The distance between the dotted lines equals the contrast in panel E. **H)** Overlay of a generalized additive model fitted to the data with pointwise determined 95% confidence interval bands.

### RAGE is involved in LPS-induced sensitisation to tissue acidosis

In experiments with pH 7.4 injections, co-injection of azeliragon with LPS resulted in a nominal reduction of the AUC Pain by 47 (CI -20–114, p = 0.11), corresponding to 54% remaining pain (control 0%, LPS 100%, Fig. 4A,B). To test the secondary hypothesis 1, acid-induced pain was compared between skin spots injected with LPS vs. LPS and azeliragon. Compared to LPS alone, the addition of azeliragon reduced the AUC pain by 207 (CI 100–315, p < 0.001), which corresponds to 60% remaining pain (control 0%, LPS 100%, Fig. 4C,D). There was no evidence for an effect of resatorvid co-injected with LPS, neither in pH 7.4 injections, nor in acidic ones (p = 0.67 and 0.13). Exploratory addition of sex to the mixed model did not indicate sex-dependent differences by the intradermal injections (‘sex’*’injection type’ p = 0.12).

**Figure 4:**
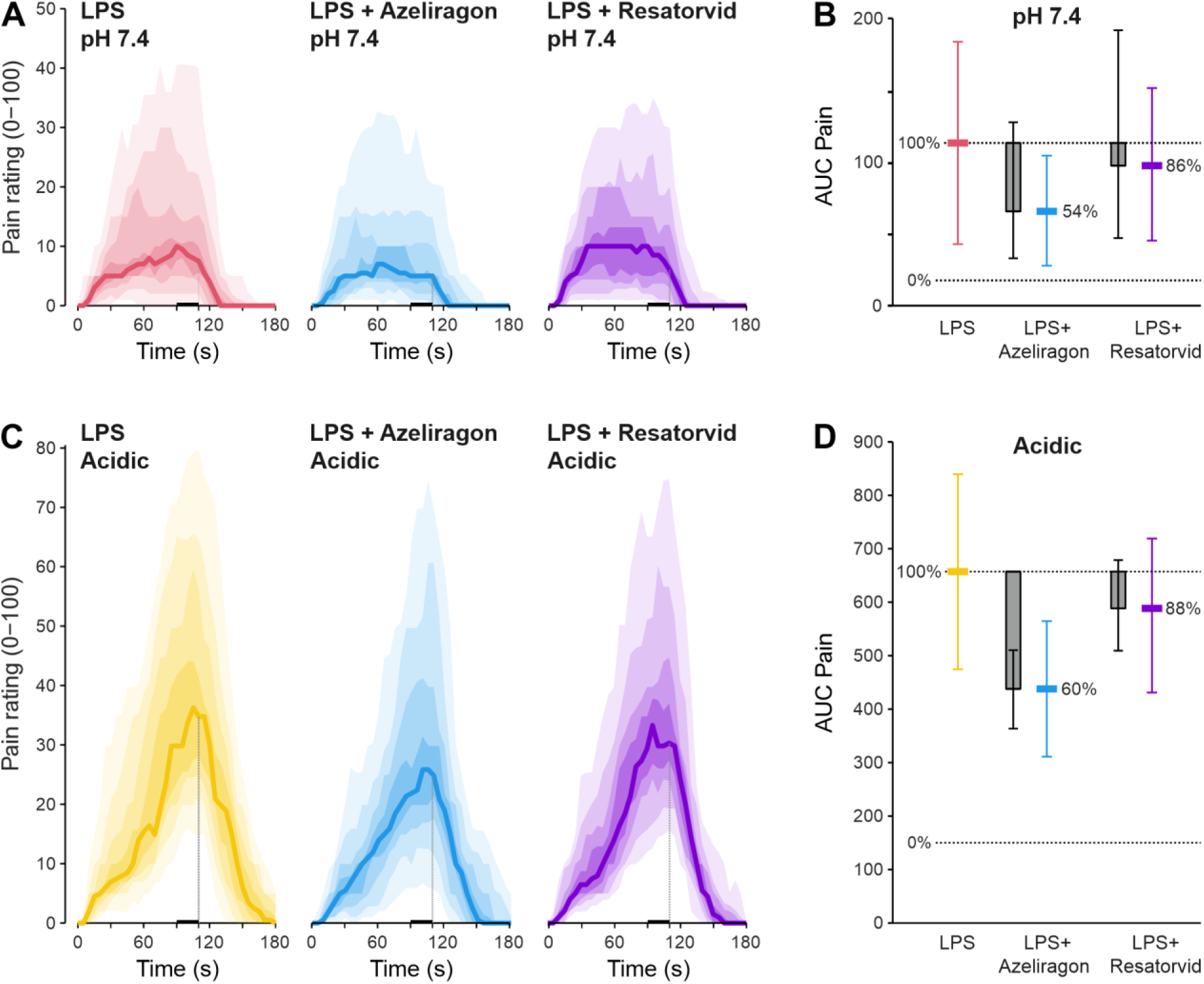
Acid hypersensitivity depends on RAGE, but not TLR4. **A)** Time courses of pain ratings for injections with constant pH 7.4, measured 4.5 h after LPS injection. The distribution is visualized with the median as a solid line and decreasing colour shades for percentiles more distant from the median (in 10% percentile steps). The dotted vertical line indicates the end of the 110 s intradermal injection. **B)** Mean estimates of pain in the period 90–110 s (AUC pain) with 95% CIs. Bars show the contrasts to only LPS for azeliragon and resatorvid with 95% CIs. A percent scale indicates the results without (0%) and with (100%) LPS pretreatment. **C)** Time courses of pain ratings for injections with increasingly acidic pH 7.2–5.4. Results without antagonists in panel A and C are identical to Figure 3. **D)** Mean estimates of pain in the period 90–110 s (AUC pain) with 95% CIs and contrast for azeliragon and resatorvid. Percentages are provided on a scale without (0%) and with (100%) LPS pretreatment.

### RAGE is the primary mediator of mechanical sensitisation by LPS

With self-closing forceps, a new mechanical pinch stimulus was established. The pain rating of 4.5 (CI 3.1-6.2) induced by mechanical pinch in naïve skin was increased to 17 (CI 13-22) in inflamed skin (Fig. 5A). For the secondary hypothesis 2, the mechanically induced pain was compared between skin spots injected with LPS alone and spots coinjected with azeliragon and LPS. The increased pain due to mechanical pinch in inflamed skin was reduced to 55% (11.3, CI 8.6-14.9) by addition of azeliragon. In contrast, addition of resatorvid did not alter sensitivity to mechanical pinching (16.1, CI 12.4-20.8). For the new mechanical pinch stimulus, the re-test reliability of three stimuli was assessed. The three repetitive measurements had an intraclass correlation of 0.29, indicating that the three pinch ratings at the same skin site were moderately consistent. At the participant level, the repeated measurements were much more consistent (intraclass correlation 0.69), indicating that each participant responded consistently across their skin spots (Fig. S2).

**Figure 5:**
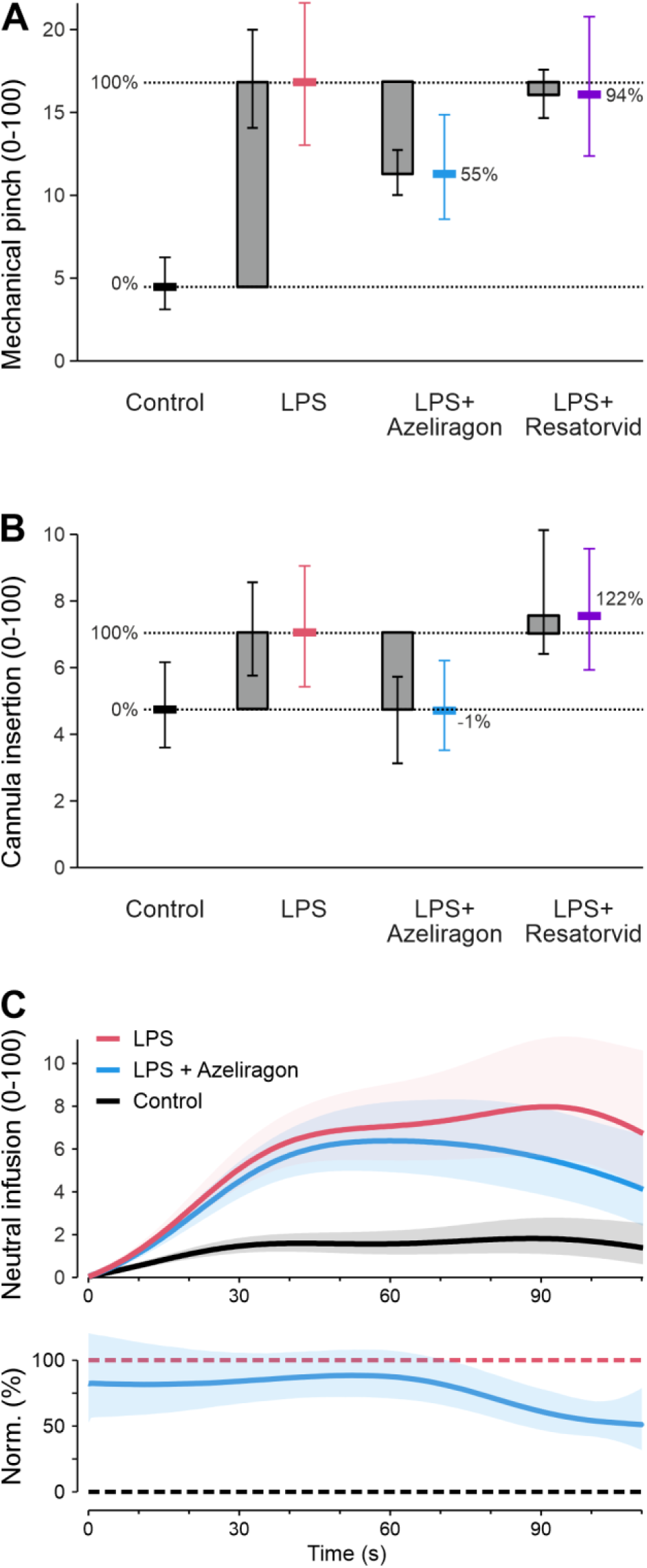
Mechanical hypersensitivity depends on RAGE, but not on TLR4. **A)** Mean estimates and 95% CIs of mechanical pain ratings, with an increase in mechanical pain 4.5 h after LPS-induced inflammation. Normalized to this sensitisation, coapplication of azeliragon with LPS reduced the mechanical pain to 55%. Resatorvid coapplication with LPS did not alter mechanical pain. Percentages of mean estimates are provided on a scale without (0%) and with (100%) LPS pretreatment. **B)** Mean estimates of cannula insertion pain ratings, again with an increase in mechanical pain 4.5 h after LPS-induced inflammation. Normalized to this sensitisation, coapplication of azeliragon with LPS reduced the mechanical pain to -1%. Resatorvid coapplication with LPS did not alter mechanical pain. **C)** Generalized additive model for the experimental groups with pH 7.4 intradermal injection. Toward the end of the injection, there is a difference due to azeliragon pretreatment, which is more apparent when the ratings of pH 7.4 injections are normalized to control (0%) and LPS (100%, bottom panel). Resatorvid had no relevant effect and was omitted for readability. Error bands are pointwise determined 95% confidence intervals.

**Figure S2.**
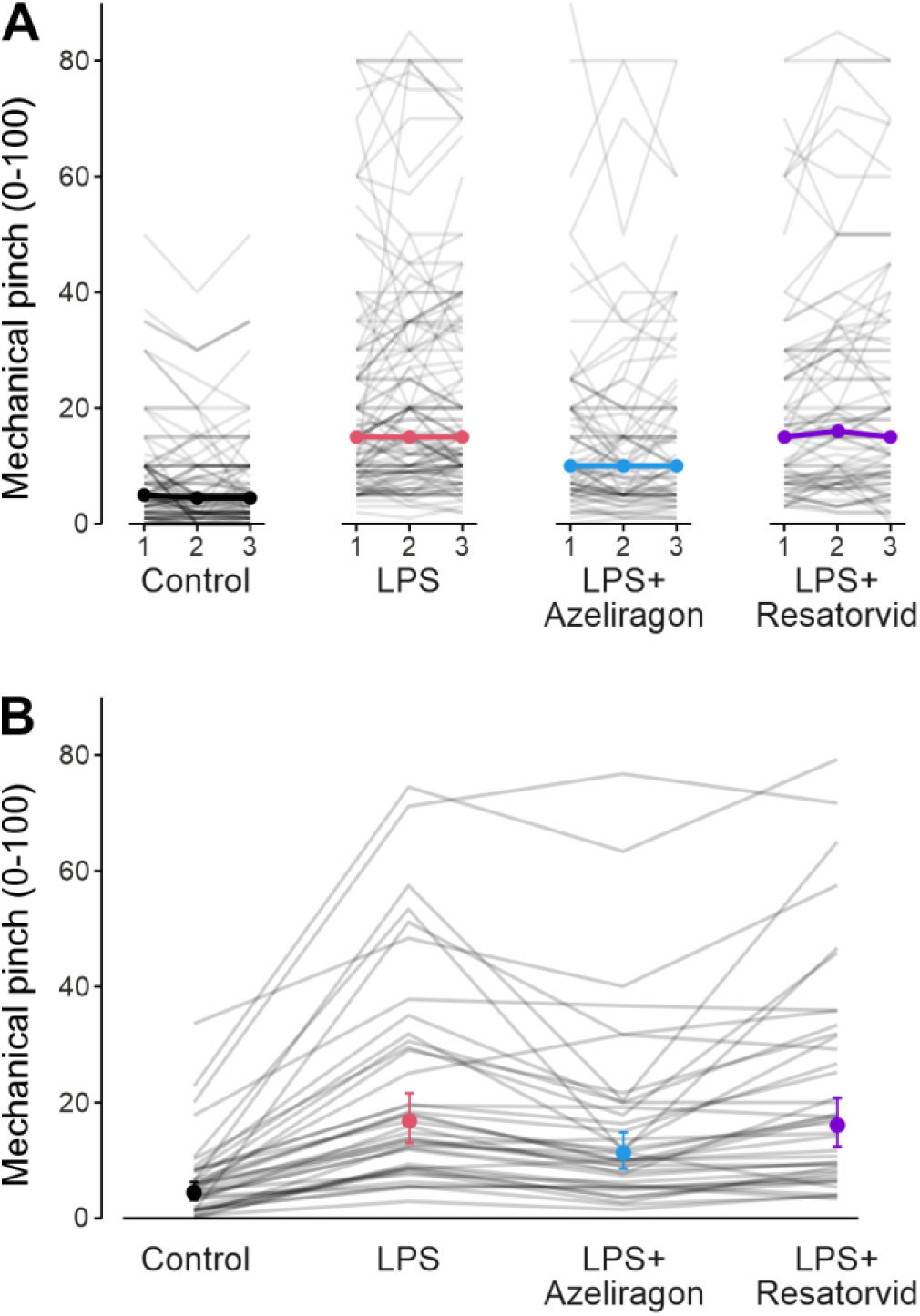
**A)** Ratings for the three subsequent pinches for all 40 subjects. The intraclass correlation coefficient, quantifying the consistency of mechanical pinch pain ratings within the same spot, was 0.29. Black lines indicate the median rating. **B)** Per subject mechanical pinch pain ratings for the four different injection types, applied 4.5 h before testing. The intraclass correlations at the subject level was 0.69, indicating the ratings are a largely stable individual trait. Mean estimates and 95% CIs are overlayed.

Cannula insertion pain is also a mechanically induced pain, and results are in line with mechanical pinch. LPS elevated the cannula insertion pain from 4.7 (CI 3.6–6.2) in naïve skin to 7 (CI 5.4–9.0) in inflamed skin (Fig. 5B). The increased cannula insertion pain in inflamed skin was abolished by addition of azeliragon, but unaffected by addition of resatorvid (4.7, CI 3.6–6.2 and 7.6, CI 5.9–9.6, respectively). The pH 7.4 injections generate a slight local distension, which caused substantially more pain after induction of inflammation by LPS (Fig. 5C). A generalized additive model was used to quantify the effect of azeliragon, which reduced the LPS-elevated pain toward the end of an injection. Resatorvid had no relevant effect.

Exploratory addition of sex to the mixed model did not indicate sex-dependent differences in pain due to mechanical pinch (‘sex’*’injection type’ p = 0.17) or due to cannula insertion into the skin (‘sex’*’injection type’ p = 0.29).

### TRPV1 mediates acid-induced pain in LPS-induced inflammation

As exploratory analysis, the contribution of TRPV1 to acid-induced pain in inflammation was investigated, considering that this is the principal detector in naïve human skin^16^.

The present study confirms the prior results, with TRPV1 antagonist BCTC largely reducing the acid-induced pain (-123, CI -189– -57, Fig. 6A-C). Also in LPS-induced inflammation, the majority of the reported pain was dependent on TRPV1 (-393.2, CI -538.0 – -248.4, Fig. 6G-I). The fraction of acid-induced pain not inhibited by BCTC was 11% (bootstrapped CI -1–31%) in naïve skin and 29% (bootstrapped CI 11-51%) in inflamed skin (p = 0.16, Fig. S3A).

**Figure 6:**
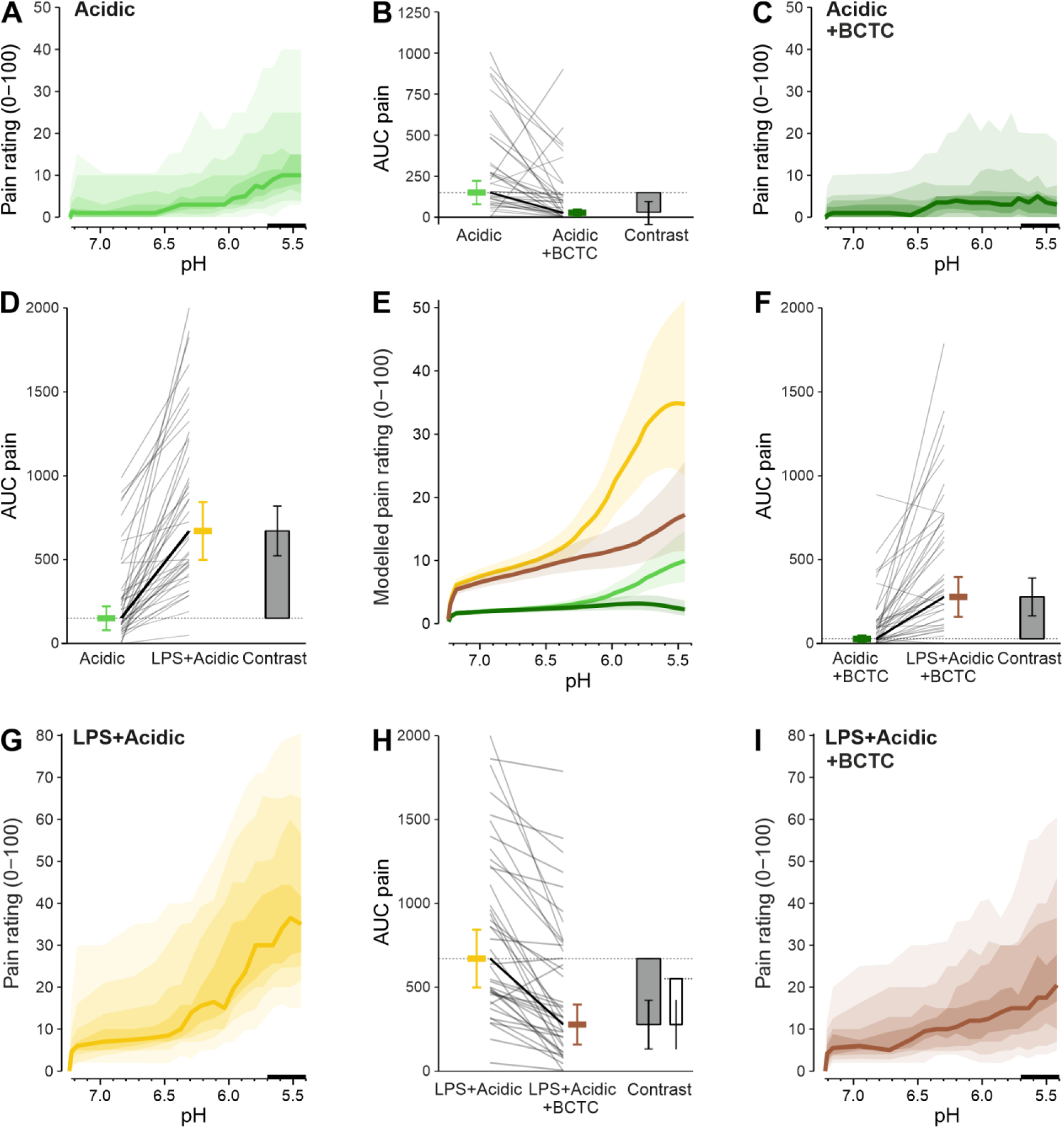
TRPV1-dependence of acid-sensing in inflammation. **A,C,G,I)** PH-Dependence of the reported pain, the distribution is visualized with the median as a solid line and decreasing grey or colour shades for percentiles more distant from the median (in 10% percentile steps, as indicated by the scale bar). This is based on the 110 s period of increasingly acidic injection, the time-dependent ratings are provided in Fig S3. Data in panel A and G are identical to Figure 3. **B,D,F,H)** Mean estimate for the AUC pain, calculated for the range of pH 5.7–5.4 (period 90–110 s, marked by a bar on the x-axis), spaghetti plot and contrast between the two groups with adjacent diagrams. Panel H also visualizes the contrast (open bar) for the prespecified hypothesis that the inhibition of acid-induced pain by adding BCTC to the injection solution is greater after LPS injection than after control injection. The distance between the dotted lines equals the contrast in panel B. **E)** Overlay of a generalized additive model fitted to the data with a 95% confidence interval.

Based on the generalized additive model used in the preceding analyses, posterior sampling was employed to estimate the pH required to produce a given difference in pain rating between two injection types. The pH required to increase the rating by e.g. 1 on a 0-100 scale was 6.5 in non-inflamed control spots, but this shifted to pH 7.2 in inflamed spots (Fig. S3B,C). Although TRPV1 explains the majority of the acid-induced pain, it is not the most sensitive pH sensor in the presented model. In naïve skin, lowering the pH below 6.5 raised the pain by 1 unit, whereas a reduction of pain by 1 unit due to TRPV1 blockade only occured below pH 6.1 (Fig. S3C,D). Consequently, the acid-induced pain observed between pH 6.5 and 6.1 cannot be attributed to TRPV1 and therefore remains unexplained.

Similarly, in inflamed skin, the pH threshold defined as an increase of 1 pain unit is about 7.2 or higher (with the range above not well addressed by the chosen pH). TRPV1, however, only explained a difference of 1 pain unit below pH 7.0 (Fig. S3C,D). This means that what has been observed in non-inflamed skin also applies to inflammation: there is a pH range in which protons already activate nociceptors, but independent of TRPV1.

Direct overlay illustrates the difference between the RAGE-dependent general reduction of inflammation and the TRPV1 component identified by BCTC (Fig. S4). Blocking RAGE reverses about half of the LPS-induced hyperalgesia across the entire examined pH range. In contrast, BCTC attenuated pain below the respective threshold of TRPV1, but reached a greater degree of pain inhibition than RAGE blockade. The full time course of pain ratings for the injection-based experiments are provided for all participants (Fig. S5).

None of the participants reported any adverse effects or taking anti-inflammatory drugs related to the study within 72 hours.

**Figure S3.**
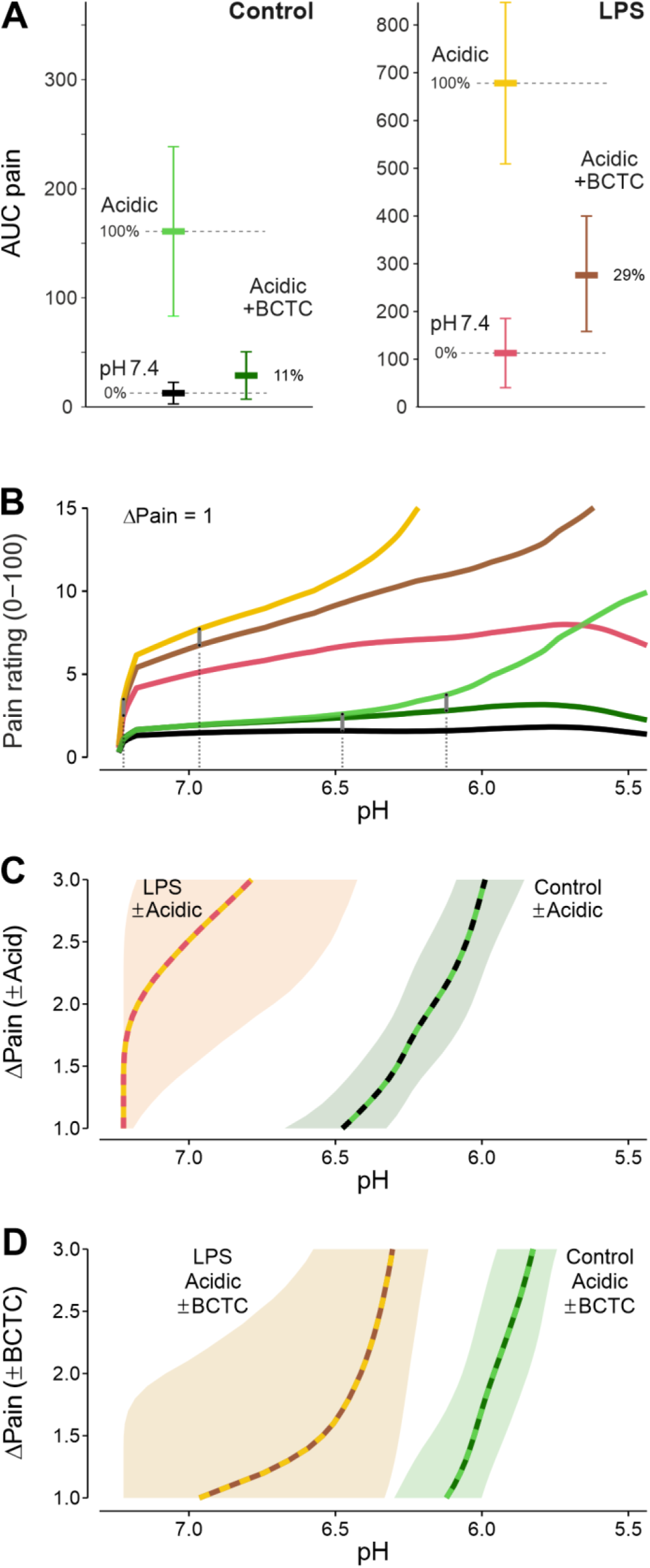
TRPV1-dependence of acid-induced pain in naïve and inflamed skin. **A)** Mean estimates and 95% CIs from figures 3 and 6 for direct comparison. In naïve skin, the fraction of pain remaining in the presence of BCTC is 11%, the confidence interval includes complete inhibition. At 4.5 h after induction of inflammation by LPS, but the fraction remaining in the presence of BCTC is estimated at 29% on the added scale without (0%) and with LPS pretreatment (100%). **B)** Overlay of generalized additive models to visualize the pH at which the pain rating differed by 1 unit. For the groups with pH 7.4, the time of the rating was projected onto the corresponding pH of the increasingly acidic groups. **C)** Without inflammation, increasingly acidic compared to pH 7.4 stimulation increased the rating by about 1 at a pH of 6.5 (dashed green-black, as green is used for acid and black for the control). In LPS-induced inflammation, a difference of 1 was present from pH 7.2. Also for higher increases of the pain rating, a 0.7–1.0 higher pH was sufficient in inflammation for a similar increase in pain compared to non-inflamed control. Data are visualized from 1 as a meaningful difference up to a delta of 3, which is available for all posterior samples. For this range of ΔPain values, the pointwise determined 95% confidence interval bands for the pH levels are provided**. D)** Without inflammation, the TRPV1-dependent acidic pain is indicated by comparing the groups with and without BCTC, which show a rating difference of about 1 at a pH of 6.1. In LPS-induced inflammation, the same difference of 1 was present pH 7.0, demonstrating inflammatory TRPV1 sensitisation.

**Figure S4.**
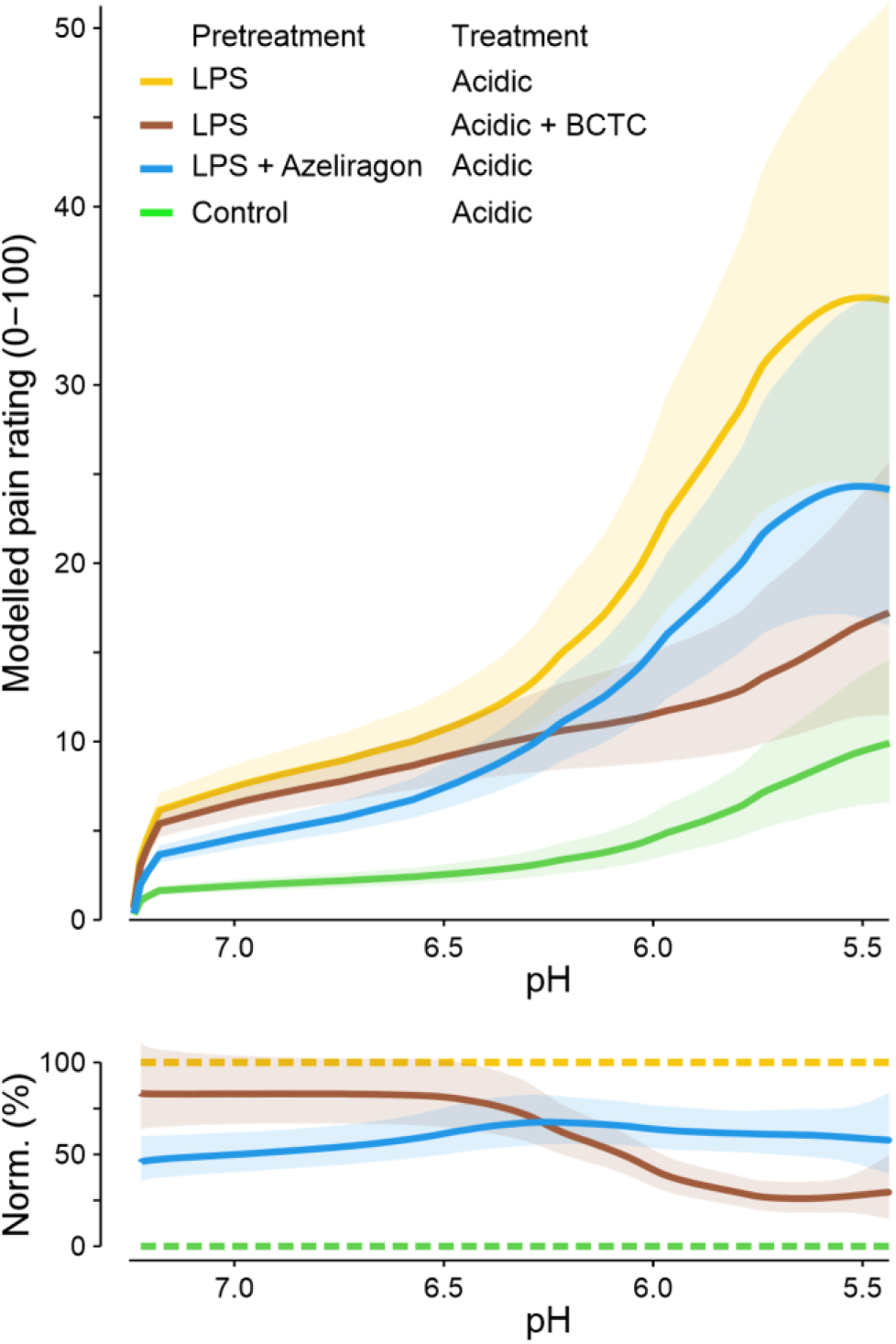
TRPV1 and RAGE contribute differently to inflammatory sensitisation. Pain ratings were modelled by a zero-anchored general additive model, which provided 95% CI bands of the mean estimates. The pH-dependence of pain in inflamed skin with vs. without azeliragon pretreatment shows a difference throughout the whole pH range. This is apparent in the bottom panel, where the ratings are normalized to acid-induced pain 4.5 h after control (0%) and LPS (100%), suggesting a rather uniform action of azeliragon against inflammatory sensitisation. In contrast, the addition of BCTC only acts below a pH threshold against the sensitized TRPV1 receptor.

**Figure S5.**
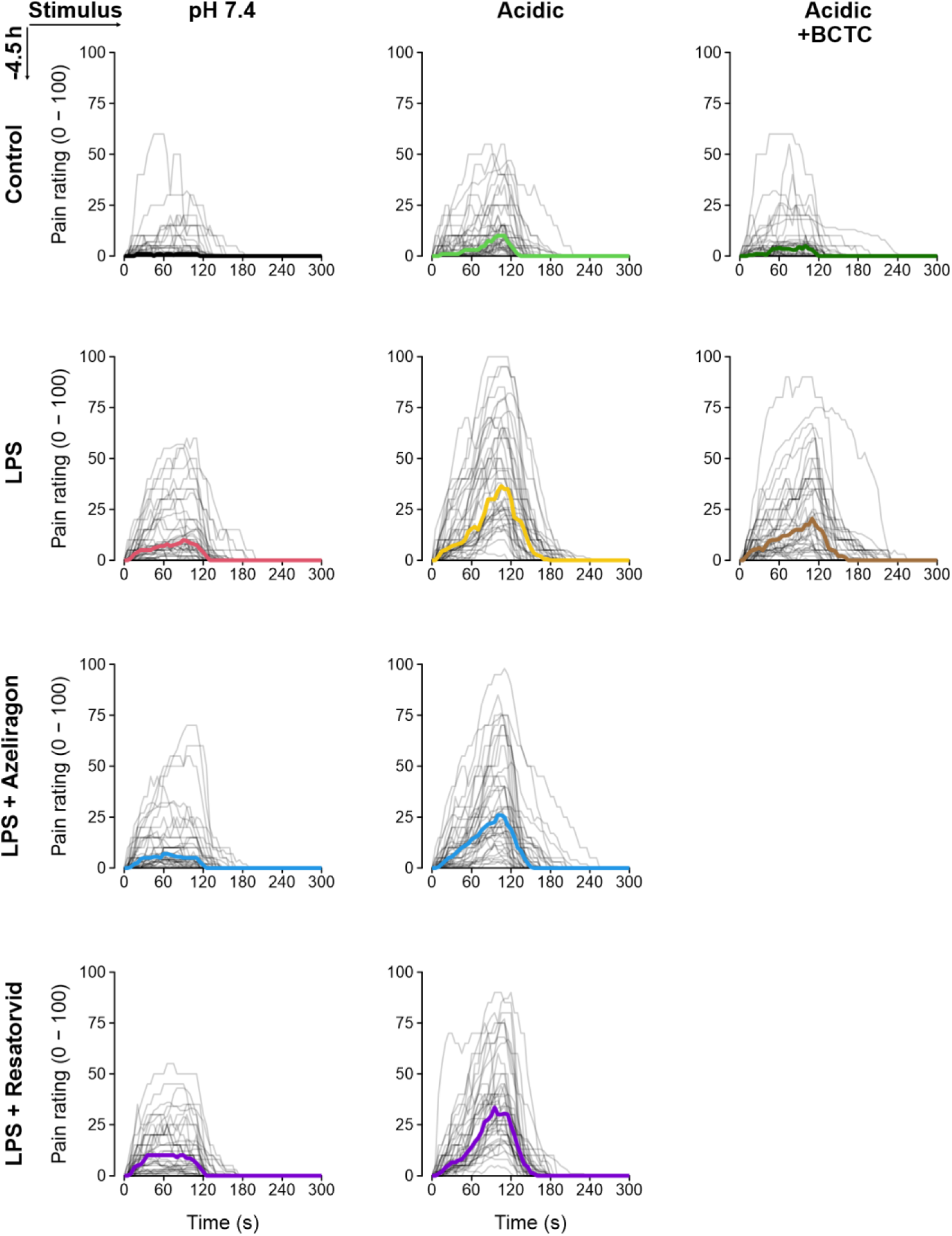
Raw data and medians for all injection pain ratings of the main study. Full time courses of pain ratings of all experimental groups. All injections lasted 110 s. The individual traces of the 40 participants are provided in light grey, the median as a solid line. Rows have the same pretreatment 4.5 h before testing, columns have the same pain-inducing injection.

## Discussion

The time course of hyperaemia and pain sensitisation was quantified in an LPS human skin inflammation pain model. In this model, LPS-induced hyperaemia and pain sensitisation depend on RAGE, but not TLR4.

The most common bacteria causing skin and soft tissue infections are by far gram-positive Staphylococcus aureus and Streptococcus pyogenes, and there are experimental models for gram-positive infection, e.g. by intradermal injection of UV-killed Streptococcus pneumoniae^22^. Nevertheless, gram-negative bacteria, like Enterobactarales and Pseudomonas aeruginosa, are prevalent in up to 14%^23,24^, and mixed infections in up to 24% of culture-positive cases^24,25^. LPS makes up several percent of the total dry mass of gram-negative bacteria^26^. Being highly proinflammatory, it is established in cellular models^27^, animal models^28^ and also as a endotoxemia model in healthy subjects^29–31^. Comparison with other established inflammatory pain models, e.g. intraplantar injection of carrageenan in rodents, shows important differences. There, signs of inflammation such as increased paw thickness and temperature peak at 2.5–4 h post-injection in rats, just like hyperaemia does in the human LPS model. However, thermal and mechanical hypersensitivity mirror the time course of hyperaemia in the carrageenan model^32^. This is clearly different in the presented human LPS model, where mechanical hypersensitivity lag behind the hyperaemia, and hypersensitivity to acid seems to be decoupled from hyperaemia.

TLR4 is the established main target of LPS both in cellular^33,34^ and animal models^9,35,36^, well summarized in a review^37^. With TLR4 and its co-factors CD14 and MD2 being present, the TLR4-dependent pathway has been shown to be intact in DRG neurons^38^. Exposure to LPS 0.02 µg/ml for 5 minutes raised intracellular calcium levels of DRG neurons and also sensitized capsaicin responses^8,39^. In addition to TLR4, RAGE was suggested to bind LPS directly^11^, leaving the share of both TLR4 and RAGE and the relationship between them open^40^, and more so in human disease. TLR4 activation triggers gene transcription of proinflammatory mediators, which takes rather hours than minutes. RAGE-dependent direct sensitization of capsaicin responses in DRG neurons has been observed after 24 h of LPS incubation^10^. Although direct effects have been reported for LPS 1 µg/ml and above^41^, there is little evidence for direct sensitization of DRG neurons by LPS in a pathophysiologically relevant concentration. Consequently, a delayed response involving other cells, such as immune cells appears to be essential. In the present study, the inhibition of RAGE largely blocked hyperaemia. The latter is an inflammatory response that is mostly independent of sensory neurons. It remains unclear if direct binding of LPS to RAGE is the crucial step, or if RAGE is involved further downstream in an inflammatory cascade. Nuclear factor High-mobility group box 1 (HMGB1), which can also be actively secreted or passively released by activated immune cells, like monocytes/macrophages^42,43^, or dendritic cells^44^, is a classical RAGE ligand^45^. Serum HMGB1 levels were elevated in endotoxin challenged mice and sepsis patients and administration of anti-HMGB1 antibodies attenuated endotoxin lethality in mice^43^.

Unexpectedly, there is no contribution of TLR4 in the LPS skin inflammation model, not to hyperaemia, not to mechanical pain hypersensitivity and not to acid-induced pain hypersensitivity. However, there is a substantial RAGE-dependent fraction, which varies across the considered readouts. Hyperaemia is almost completely RAGE-dependent, but less than half of acid-induced pain. The mechanism of LPS-induced sensitisation leading to the remaining fraction of mechanical and acid-induced pain is unclear. E.g. a role of caspase-4/5 and a downstream pathway leading to NLPR3 inflammasome activation and IL-1beta/IL-18 secretion might be considered^46^.

For pharmacological inhibition of TLR4 and RAGE, substances that reached at least clinical phase II trials were considered, as this implies comprehensive preclinical toxicology studies and a phase I safety study. Paridiprubart (NI-0101), an anti-TLR4 monoclonal antibody, has reached clinical phase II trials. Eritoran (E5564), a synthetic lipid A analogue that antagonizes the TLR4/MD2 complex, failed to show efficacy in phase III, just like resatorvid. Resatorvid was chosen because it is well established in preclinical research and because it is a small molecule. At present, azeliragon (TTP-488/PF-04494700) is the only RAGE antagonist having reached phase III clinical trials.

When considering the translation to other body sites, exposure to LPS ranges from almost none in the lungs to a massive commensal LPS load in the gut. Complementary to this, there is a strong inflammatory response to LPS in the lung, but a state of controlled tolerance in the gut. The skin represents an intermediate environment, with its commensal flora rich in Gram-negative bacteria. However, compared to mucosal surfaces, the role of skin-specific cell types need to be considered.

Keratinocytes can respond to LPS, releasing neutrophil attractant IL-8, monocyte and macrophage attractant MCP-1 and CCL2, IL-6 and TGF-beta^47,48^. In epidermal Langerhans cells, the combination of TNF-α/IL-1β and LPS supra-additively drives development of a mature immunogenic state^49^. Also human dermal fibroblasts contribute to the LPS response by a non-canonical pathway^50^. Therefore, body site-specific cell types and signalling pathways could allow a context-sensitive response and require validating findings from skin inflammation at other sites.

With respect to the relative contribution of TRPV1 to acid-induced pain, in our prior study investigating the contribution of different ion channels to acid-induced pain, there was no inhibition by ASIC antagonist amiloride and TRPA1 antagonist A-967079. In contrast, application of TRPV1 antagonist BCTC largely reduced pain (70%, CI 53-84%)^16^. The novel aspect of this study is its examination of TRPV1’s role in inflammatory conditions. Unsurprisingly, TRPV1 still contributes, and this is still the majority. Although there is no statistical difference in the TRPV1-dependent fraction in naïve and inflamed human skin, the estimated TRPV1-independent component is 11% in naïve skin and 29% in inflamed skin. Nevertheless, inhibiting TRPV1 has the potential to reduce pain in all pathophysiological conditions involving tissue acidosis.

These findings raise the question of how inflammation affects the pH sensitivity and the activation threshold of TRPV1. TRPV1 has a threshold for activation at about pH 6.0^51,52^, and considering synergistic activation, pH 6.4 clearly sensitizes against capsaicin or temperature activation^51^. A key feature of TRPV1 is the extensive, rapid and rapidly reversible sensitisation^53–55^. Also pH sensitivity was demonstrated to be increased by e.g. ATP, in ischemia, or after induction of inflammation in animals^17,56,57^. However, the extent of a threshold shift was not thoroughly quantified in vitro. Unfortunately, reported pH levels at which there is an increased pH response were not designed to detect a threshold, had widely spaced pH levels and lacked the gradual stimulus increase, all contributing to an underestimation of the threshold. The pain rating differences in the generalized additive model allowed quantifying the pH threshold shift in the LPS-induced human skin inflammation model. For example, the pain rating increased by one point on the 0–100 scale at a pH of 6.5 compared to pH 7.4 in naïve skin, but already at a pH of 7.2 in inflamed skin. The shift in pH sensitivity occurs for the TRPV1-dependent as well as for the TRPV1-independent fraction of pain. The observed level of sensitisation, even exceeding prior cellular data, has a pathophysiological relevance, so that frequently occurring and mild reductions of pH can be expected to contribute to pain in humans.

It is equally important to understand how inflammation modulates mechanical sensitivity, the second key sensory modality in this model. Mechanical sensitivity, assessed by the quantitative sensory testing battery^58^ ^59^ contains non-noxious mechanical stimulation, but also mechanical pain thresholds with focal as well as blunt pressure stimulation, and finally assess suprathreshold mechanical pain sensitivity with pin pricks^58^. The assessment of suprathreshold pain was the main goal of the present study. However, the strongest 512 mN pinprick stimulator of the quantitative sensory testing protocol generates low pain ratings in naïve skin and also did not reflect the marked mechanical hypersensitivity due to LPS-induced inflammation well in pilot experiments. In particular, lateral shear stress, e.g. even a mild stroke with a fingernail across the site generated a pronounced difference in sensation between inflamed and naïve skin. The search for an adequate stimulus resulted in a pinch, delivered by self-closing forceps used to clamp structures in surgery. The stimulus generates no or marginal pain without inflammation in the majority of subjects, but is clearly painful in inflammation. To improve precision, this was measured three times, a higher number of tests was avoided considering wind-up paradigms. The sequential ratings do suggest neither a sensitisation nor a desensitisation across the triplicate measurement. Azeliragon reduced the pain induced by pH 7.4 injection, which can only be attributed to mechanical phenomena. The needle insertion pain was analysed as an exploratory outcome. Nevertheless, it provided convergent evidence for the mechanical pinch results, as the needle insertion pain was also sensitized by LPS, and reduced by azeliragon but not by resatorvid.

Inflammatory sensitisation to heat is well established from cellular assays to human models, even including UV-based inflammation^60^. The pilot study was designed to measure time course, but also amplitude and variability of the expected heat sensitisation, but surprised with a lack of heat sensitisation in LPS-induced inflammation. A TRPV1 contribution to this threshold was expected, due to the observed shift in TRPV1 sensitivity for acid detection. A possible explanation is a modality-specific sensitisation, which was described to be largely dependent on a threonine at position 704 for sensitisation to heat, but sensitisation to acid relies on a serine at position 800^61^. Position 704 is mainly phosphorylated by PKCβII^62^, in contrast to 501 and 800, which are mainly phosphorylated by PKCε ^63^.

Considering the upstream signalling cascade, both PKC isoforms are activated by DAG, but the conventional PKCβII also requires a calcium elevation^64^. A limited calcium signal would therefore be one possible explanation of the absence of heat sensitisation. Regulation of expression of protein kinases and an altered signalling e.g. by cytokines in the inflammatory response should also be considered. The area of hyperaemia detected by laser speckle imaging 4.5 h after LPS injection corresponds to a circle with a diameter of 18 mm. The thermode stimulates 7.4 x 24.2 mm, so also surrounding skin, but in case of substantial sensitisation, this is expected to dominate the heat sensitivity.

Considering safety of the presented model, the doses of LPS are low compared to the established intravenous LPS challenges with a dose range 0.06 - 4 ng/kg, reaching total systemic doses of up to 300 ng. Intradermally, 5-15 ng LPS were applied per injection with total doses per subject of up to 45 ng, leading to small-diameter inflammatory lesions that completely subside within 48 h. Importantly, in these studies, no systemic signs or symptoms were observed ^7,65,66^, which also applies to our study. LPS is slowly redistributed from an injection spot. It takes several days until 50% are removed. When it has reached the circulation, it is removed within minutes by the liver^67^. Therefore, after a peripheral LPS injection at ‘no point was more than 1% of the injected dose found in the plasma^68^. Both reproducibility and safety benefits from the highly standardized LPS production. Despite only a few investigations, the topical LPS challenge can be considered an established and safe model of inflammation.

### Limitations of the study

We aimed to provide a complete time course of the LPS-induced inflammation. The observation interval up to 50 h after LPS injection was based on pilot experiments, and with the assumption of a limited difference between the time course of hyperaemia and pain hypersensitivity. In hindsight, the observation interval falls short describing the complete time course of hypersensitivity until complete recovery. Although there were no signs for residual inflammation after 72 h in pilot experiments, the respective time has not been acquired in the pilot study. Azeliragon was used at a relatively high local concentration (100 µM) to ensure robust RAGE blockade. This concentration was chosen to largely inhibit RAGE for a considerable period. However, effects of other targets cannot be excluded. E.g. p21-activated kinase 1 is inhibited by azeliragon^69^, and has a role in pain models^70^. The cannula insertion pain and the pain ratings in the first seconds are different between naïve and inflamed skin. This could have provided a cue, and limited the blinding of the subject. Using only a single molecular component of bacteria might not cover all aspects of a skin infection, as the model lacks other bacterial components. Nevertheless, this is outweighed by the advantage of a biologically well-defined model, triggering a reproducible degree and time-course of inflammation.

Taken together, we have established a novel human pain model based on intradermal LPS administration. RAGE, but not TLR4, is crucial for mediating the resulting inflammatory hyperaemia and pain hypersensitivity, highlighting RAGE as a promising therapeutic target for inflammatory pain.

## Methods

Ethical approval was granted by the Ethics Committee of the Medical University of Vienna for investigating the time course of inflammation in a pilot study (vote 1569/2023). After completion, a study regarding the involved signalling mechanism and the contribution of TRPV1 was designed and approved (vote 1692/2024). Following comprehensive instruction regarding the nature, significance, impact and risks of this study, the subjects provided written consent to participate in the study. In addition, the participants received a written information sheet in comprehensible language, explaining the nature and purpose of the study and its procedures. The participants were informed that they could withdraw their consent at any time without providing reasons. In female subjects, a pregnancy was excluded by a test prior to any study-related intervention. Completion of the study visit was financially compensated as approved by the ethical committee. Participants had insurance for participation and were contacted 72 h after completion of the study to ask whether any unanticipated adverse effects had appeared and whether anti-inflammatory drugs had to be taken. To comply with data protection laws, no protected information was digitised. Each participant is represented by a sequential number in all electronic records. The study was conducted in concordance with the principles of the ‘Declaration of Helsinki’ (as amended at the 64th WMA General Assembly, Fortaleza, Brazil, 2013) and with the laws and regulations of Austria. Only appropriately trained personnel were involved in the study. Before inclusion of the first subject, the main study was registered on https://www.clinicaltrials.gov (NCT07092215) on 11^th^ of June 2025. The pilot study was carried out between 20^th^ of November-21^st^ of December 2023, the main study between 17^th^ of June and 11^th^ of July 2025 at the Medical University of Vienna, Vienna, Austria.

### Experimental model of skin inflammation

In contrast to systemic LPS administration, dosed in the range 0.06–4 ng/kg^71^, local intradermal LPS application has been established with a dose of 5–15 ng per injection, leading to small-diameter inflammatory lesions that completely subside within 48 h^65,66^. A dose of 5 ng LPS per injection was validated in own pilot experiments, also e.g. 10-fold lower concentrations caused hyperaemia and hypersensitivity. The pilot study had 9 injections containing LPS, the main study had 7 injections, resulting in a total dose of 45 and 35 ng per subject. A volume of 50 µl was superficially injected into premarked skin spots using a 30G syringe (BD, Micro-Fine U-100, 0.3 x 8 mm).

### Quantification of hyperaemia

The degree of hyperaemia was quantified for both forearms by using the laser speckle contrast imager moorFLPI-2 (Moor Instruments, Axminster, United Kingdom) and the moorFLPI-2 Measurement V3.0 software. The forearm was placed on a black neoprene mat and the built-in aiming laser was used to standardize the distance between camera and skin surface. The subject was asked not to move the forearm for 10 s to acquire a colour image and five speckle images sampled from 2 s each (20 ms exposure time with a 100 frames temporal filter). The least blurred laser speckle image was extracted as a 8-bit monochrome image with a background threshold of 10 (moorFLPI Review Software V6.0, Moor Instruments, Axminster, United Kingdom). ImageJ v1.54p was used for further analysis. The image was binarized to generate regions of interest marking inflamed skin spots by three ‘erode’, followed by eight ‘dilate’ operations. Surrounding reference regions were generated by ten further ‘dilate’ operations. Pixel areas and mean grey values were then used to calculate a reference-subtracted volume integral (Δblood flow) for every skin spot. Non-inflamed skin spots were barely distinguishable from the background in the speckle image. Thus, using the colour image, a circle with a 3.2 mm radius around the injection spot (pre-marked by a pen) accompanied by a surrounding reference circle with a 4.9 mm radius was used to calculate the Δblood flow of non-inflamed skin spots.

### Subjective pain rating paradigms

Participants were first familiarized with the respective pain stimuli at native skin spots. For all rated stimuli, perceived pain was reported on a 0–100 scale, whereby 0 represents no pain, and 100 represents the worst imaginable pain. The heat pain threshold was determined by instructing participants to press a button when the increasing thermode temperature was first perceived as painful.

### Heat pain threshold

The heat pain threshold was assessed according to the standardized protocol for quantitative sensory testing^58^. The heat pain threshold was determined with a T11 probe operated by a TCSII thermal stimulator (QST.Lab, Strasbourg, France). Skin spots were stimulated with one activated zone (7.4 x 24.2 mm). Starting from 30°C, a predefined temperature ramp with +1°C/s was applied. Each skin spot was consecutively tested three times.

### Mechanical pinch

A mechanical pinch stimulus was realized using cross-over forceps (Universalkreuzpinzette, 92 94 91, Knipex, Wuppertal, Germany), customized with a 3D-printed inset to have a maximal opening diameter of 6 mm, which fits the total area of erythema arising from the inflammation model used. For stimulation, the levers were fully opened by squeezing, gently placed onto the test site, and closed by releasing. At the resulting opening diameter of about 3 mm, the forceps apply a standardized pinch force of 2.560 mN. Forceps were removed after ∼1s and intensity of the perceived pain was rated by the subject. Each skin spot was tested three times in random order.

### Experimental model of acid-induced pain

To apply a defined nonhazardous intradermal acidic stimulus, previously established injection-based pain models^16,21^ were further improved to expose human skin to a largely linear pH ramp. Before each injection a pH 7.4 and a pH 5.0 solution were loaded in 5 ml syringes (Injekt Luer Lock Solo) and connected to extension lines (Original Perfusor Line, 50 cm, both from B.Braun, Melsungen, Germany). The extension lines were connected to a 30 G cannula by a 3-way stopcock (Sterican, 0.30 x 12 mm and Discofix 3SC, both from B.Braun, Melsungen, Germany). The syringes were inserted into two programmable pumps (World precision instruments, Sarasota, FL). The pH 5.0 syringe was flushed until the fluid reached the cannula tip, followed by flushing the 7.4 syringe for 5 s. For pH measurements, the fluid of three runs was collected in fractions of 5 s each (i.e. 22 fractions for a 110 s protocol) and pH was measured at room temperature using a calibrated pH microelectrode (Biotrode, 913 pH Meter, Metrohm, Herisau, Switzerland). This procedure was repeated three times. Over a period of 110 s, the two programmable pumps stepwise adapt their injection rates to generate a linear decrease from a pH of 7.24 to a pH of 5.44. The added injection rate from both pumps is at a constant 30 ml/h throughout the protocol, which results in a total volume of 0.92 ml administered per injection. The cannula was intradermally inserted into the respective spot on the volar forearm of the subject and fixed with Leukoplast (BSN medical, Hamburg, Germany). Pain arising from cannula insertion was then rated as a single number from 0-100. When the insertion pain had fully subsided, the pump protocol and an analog stopwatch were started. Pain was rated every 5 s until the subject reported no pain for a period of 30 s. The pain ratings were noted on paper by the experimenter. After removing the cannula, there was an at least two minutes break before testing the next skin site.

### Applied solutions and substances

The buffered interstitial fluid adapted from Bretag^72^ was used as previously described^73^. It contained (in mmol/L) NaCl 113.8, KCl 3.5, Na_2_HPO_4_ × 2H_2_O 1.7, MgSO_2_ × 7H_2_O 0.7, sodium gluconate 9.6, glucose × 1H_2_O 5.0, sucrose 7.6, CaCl_2_ × 2H_2_O 1.5, and histidine 22.0, dissolved in ddH_2_0 (Milli-Q system, Millipore, Burlington, MA). The solutions were titrated to a pH of 7.4 or a pH of 5.0, respectively, followed by sterile filtration using a 0.2 µm filter (Filtropur S, 0.2 µm, Sarstedt, Nümbrecht, Germany). Lipopolysaccharide was acquired as a highly purified molecule obtained from Escherichia coli Type O113 from List Biological Laboratories Inc. (Campbell CA, USA)^74^. LPS was stored in a stock solution of 1 mg/ml in ddH_2_0. For the injection solution, LPS was dissolved in interstitial fluid with a pH of 7.4 to a concentration of 100 ng/ml. The RAGE antagonist azeliragon has an IC_50_ of 1 µM^75^ and was orally dosed with 5-50 mg per day in three phase III clinical trials (NCT02080364, NCT02916056 and NCT05815485). Here, azeliragon (TargetMol, Boston, MA, purity 96.7%) was used at a concentration of 100 µM, which was diluted from a 10 mM stock solution in DMSO. The total azeliragon dose per subject in the present study was 5.3 µg, corresponding to ∼0.1% of the daily oral dose used in the phase III clinical trial. TLR4 antagonist resatorvid (IC_50_: 110 nM)^76^ was investigated in two phase III clinical trials (NCT00143611, NCT00633477; 2.4 mg/kg/d s.c.). Here, resatorvid (MedChemExpress, Monmouth Junction, NJ, purity 98.3%) was used at a concentration of 10 µM, which was diluted from a 1 mM stock solution in DMSO. The total dose of resatorvid was 0.4 µg per subject, corresponding to ∼0.0002% of the estimated daily s.c. dose used in the clinical trials. All pre-injections at time point 1 in the main study contained 1% DMSO.

TRPV1 antagonist BCTC (Cayman Chemical Company, purity 100%) was added from a 1 mM stock solution in DMSO. BCTC has an IC_50_ of 0.7 nM for acid-induced activation of human TRPV1^77^, and was previously used in injection-based pain studies at a final concentration of 1 µM^16,78,79^. In order to compensate for adsorption of the lipophilic BCTC to the plastic tubing in the acid-induced pain model, the needle outflow from a regular protocol was collected and analysed by high-performance liquid chromatography (HPLC). BCTC was quantified using RP-C18 columns (150 × 3.0 mm, 2.5 μM; Kinetex, Phenomenex, Torrance, CA), in a Dionex Ultimate 3000 UHPLC (ThermoFisher, Waltham, MA). The mobile phase was a gradient from solvent A (water with 0.1% trifluoroacetic acid) towards solvent B (10% water, 0.1% trifluoroacetic acid, 90% acetonitrile, VWR Chemicals, Fountenay-sous-Bois, France) with a flow rate of 0.3 mL/minute. The gradient started at 95% A/5% B and was linearly raised to 80% B over a period of 25 minutes. Absorbance was monitored at 254 nm. Results indicated a substance loss of approximately 1/3 due to adsorption to tubing, which was compensated for by adjusting the concentration of the prepared solution by a factor of 1.5. The HPLC-validated finally applied concentration was 102% of the target concentration. All injections at time point 2 in the main study contained DMSO 0.15%. All antagonist concentrations were chosen considering local dilution and redistribution.

Solutions used for pre-injections were aliquoted in 2 ml glass screw top vials with pierceable caps (polytetrafluorethylen/silicon, Agilent, Santa Clara, CA). Solutions used for the acid-induced pain model injections were aliquoted in 12 ml glass screw top vials with pierceable caps (ND20, Butyl/PTFE, DWK Life Sciences, Wertheim, Germany). All solutions were prepared in advance and stored at -20°C until the experimentation day. Each injection in the acid-induced pain model required two separate syringes; therefore, vial pairs were prepared and labeled ‘7.4’ and ‘5.0’, respectively. For non-acidic control injections, both vials contained pH 7.4 solutions, although the ‘5.0’ label was retained to blind the experimenter.

#### Pilot study

Temporal profiling of LPS-evoked hyperaemia and hypersensitivity in human skin The objective was to describe the time courses of LPS-induced skin hyperaemia, heat pain threshold, mechanical sensitivity and acid sensitivity. This first study was designed to provide the basis for sample size calculations for the subsequent confirmatory study also and to identify which measures have a relevant difference to the baseline within the overserved 50 h period. The pilot study was prespecified to not investigate a hypothesis.

### Hypothesis-testing study: RAGE versus LPS-induced cutaneous sensitization

The objective of this study was to elucidate the mechanisms of LPS-induced hypersensitivity. This objective comprises three independent, pre-specified research questions. The mediation of proinflammatory effects of LPS via TLR4 has long been established. The question remained, whether parts of the proinflammatory effects of LPS are also mediated through an alternative RAGE-dependent signalling pathway.

#### Primary hypothesis

‘The degree of hyperaemia is different in LPS-injected skin spots compared to LPS-injected skin spots where azeliragon is co-injected’.

#### Secondary hypothesis 1

‘Acid-induced pain is different in LPS-injected skin spots compared to LPS-injected skin spots where azeliragon is co-injected’.

#### Secondary hypothesis 2

‘Mechanical pinch-induced pain is different in LPS-injected skin spots compared to LPS-injected skin spots where azeliragon is co-injected’.

As a reference, the contribution of TLR4 was measured using the antagonist resatorvid. In addition, there were two prespecified, yet exploratory research questions. The first question addressed the sensitization in the LPS inflammation model with the hypothesis ‘The pain induced by acid in addition to mechanically induced pain (by injection into the skin) is greater after LPS injection than after control injection’. This assessed the pain induced by injection of increasingly acidic (pH = 7.2–5.4) or neutral (pH = 7.4) solutions, allowing differentiation between purely mechanical hypersensitivity and additional hypersensitivity to acid. The second question addressed the role of TRPV1 in acid-induced pain in the LPS inflammation model with the hypothesis: ‘The inhibition of acid-induced pain by adding BCTC to the injection solution is different after LPS injection than after control injection’. This assessed the portion of acid-induced pain attributable to TRPV1 activation changes due to LPS-induced inflammation.

### Inclusion and exclusion criteria

For both studies, the inclusion criteria were an age between 18 and 70 years and full legal capacity. Exclusion criteria were participation in another study, ongoing or within the last four weeks, current medication intake except hormonal contraception, drug abuse, body temperature above 38°C, known allergic diseases, in particular asthmatic disorders and skin diseases, a sensory deficit or hematoma of unknown origin in the physical examination of the test site, or symptoms indicating respiratory tract infection. Female subjects currently breastfeeding or with a positive pregnancy test provided at the beginning of the study visit were excluded. To ensure an equal number of each sex in the study population, once one sex reached half of the target sample size, only the other sex was included.

### Sample size

Twelve individuals were included in the pilot study, which allows estimation of the variance^80^. For the hypothesis-testing study, a Williams balanced latin square design was chosen to minimize psychological carryover effects between injections and to ensure complete counterbalancing of treatment sequences. Ten different injection types were compared, therefore this design required ten unique sequence combinations and a multiple of ten for the number of participants^81^. For each participant, one of the pre-specified sequences was randomly selected and removed from the pool. For the primary hypothesis, the mean blood flow of LPS groups (D,E,F) is compared with the mean blood flow of LPS + azeliragon injected spots (G,H). The sample size calculation using pilot study data yielded a sample size of 10 participants for a relevant effect size of 35%, α = 0.05 and 1 - β = 0.8. For the secondary hypothesis 1, the acid-induced pain rating AUC Pain of LPS and LPS + azeliragon is compared (E vs. H). The sample size calculation using pilot study data yielded a sample size of 40 participants for a relevant effect size of 35%, α = 0.025 and 1 - β = 0.8. For the secondary hypothesis 2, the mean mechanical pinch pain rating of LPS groups (D,E,F) is compared with the mean mechanical pinch pain rating of LPS + azeliragon groups (G,H). The sample size calculation using pilot study data yielded a sample size of 20 participants for a relevant effect size of 35%, α = 0.025 and 1 - β = 0.8. To cope with up to 10% of participants not completing the study, ethical approval was applied for and received (EK 1692/2024) for up to 45 subjects.

### Analysis of primary hypothesis

According to the study protocol, the ten injection types (A–J, Fig. 2A) administered to each subject were grouped a priori into four groups: injections A, B and C constituted the group ‘Control’; injections D, E and F the group ‘LPS’; injections G and H the group ‘LPS + RAGE antagonist’; and injections I and J the group ‘LPS + TLR4 antagonist’. For statistical modelling, blood flow values from injections of the same group were averaged on a per-subject basis, yielding one value per group for each subject.

The primary hypothesis concerned the comparison of blood flow between ‘LPS’ and ‘LPS + RAGE antagonist’. For the inferential analysis, a linear mixed-effects model was fitted using restricted maximum likelihood as implemented in the nlme package in R. The within-subject mean of blood flow served as the dependent variable. The model included a four-level fixed factor group (‘Control’, ‘LPS’, ‘LPS + RAGE antagonist’, ‘LPS + TLR4 antagonist’) and a random intercept for each subject to account for the clustering of observations within subjects. Blood flow values were analysed on the original (untransformed) scale, with heteroscedasticity accommodated directly by allowing each condition to have its own residual variance. This was implemented using a variance-by-group structure (varIdent in nlme), which estimates a separate residual variance for each level of the group factor rather than assuming a common variance across all groups.

Estimated means and 95% Wald confidence intervals were obtained using the emmeans package (version 1.11.1) in R. The three contrasts described above were prespecified: the primary contrast (‘LPS’ vs. ‘LPS + RAGE antagonist’) corresponded to the main study hypothesis. As positive controls, two additional predefined contrasts were evaluated: ‘Control’ vs. ‘LPS’, assessing the expected increase in blood flow induced by LPS as established in the pilot study, and ‘LPS’ vs. ‘LPS + TLR4 antagonist’, based on the conventional assumption that LPS elicits inflammation through TLR4 signalling. The additional contrasts were included as confirmatory reference comparisons, therefore, no adjustment for multiple testing was applied.

### Analysis of injection-induced pain ratings

For descriptive inspection of all data involving injection-induced pain, individual time-courses of pain ratings (0–100) were plotted for each injection type (A-J). For every subject, the pain ratings induced by the intradermal injection were shown as a separate line. To summarise central tendency and variability, percentile ribbon plots were generated for each injection type, displaying nested 10–90%, 20–80%, 30–70% and 40–60% percentile bands together with the median curve. Similar plots were produced with a pH-scaled x-axis, allowing a visual assessment of pain as a function of the pH decline during injection. Collectively, these descriptive plots provided an overview of the magnitude, time course and variability of injection-induced pain across injection types before statistical modelling.

Analyses involving injection-induced pain used the area under the curve of the pain ratings as a summary outcome. For each subject and injection type, pain was recorded on a 0–100 numeric rating scale at 5 s intervals until six consecutive 0 ratings were obtained. To obtain a standardised quantitative measure of the experienced pain, the area under the curve was calculated over the predefined inclusive interval 90–110 s, corresponding to the last 20 s of the injection. Within each subject and injection type, all pain ratings within this window were numerically integrated using the trapezoidal rule based on pain (0–100) and time (seconds), resulting in one area under the curve value for each injection type per subject. This value served as the outcome variable area under the curve of pain (‘AUC Pain’) with the unit pain-seconds.

For the inferential analysis of AUC Pain, mixed-effects models were fitted with the individual AUC Pain values as the dependent variable. Injection type (ten levels, A–J) and the crossover period were included as fixed effects, and a random intercept for each subject accounted for the clustering of the ten injections within subjects.

### Analysis of secondary hypothesis 1

The secondary hypothesis 1 concerned the comparison of acid-induced pain between ‘LPS’ and ‘LPS + RAGE antagonist’. For the contrast analysis, all pairwise comparisons between injection types were obtained directly from the final mixed-effects model. On the log₁₀(AUC Pain + 1) scale, contrasts correspond to simple differences between the estimated means, resulting in ratios after back-transformation (i.e. multiplicative effects on the original AUC scale). As multiplicative results are not always intuitive for interpreting injection-related pain, additive differences on the original AUC scale were reported. These additive contrasts and their confidence intervals were computed using the regridding procedure in the emmeans framework, which applies the multivariate delta method to propagate uncertainty through the back-transformation and provides valid additive and multiplicative contrast estimates consistent with the underlying mixed-effects model.

### Analysis of secondary hypothesis 2

The secondary hypothesis 2 concerned the comparison of mechanical pinch-induced pain between ‘LPS’ and ‘LPS + RAGE antagonist’. Pain ratings elicited by the three brief mechanical pinch stimuli (pinch = 1–3) were first explored graphically. For each subject and injection type, the pain ratings of the three stimuli were averaged. Visual inspection of spaghetti plots stratified by injection type, pinch level, and grouped injection categories revealed no systematic trend across the three stimuli.

To obtain group-level estimates, the averaged pain ratings were analysed using a linear mixed-effects model with grouped injection types as fixed effect (‘Control’, ‘LPS’, ‘LPS + RAGE antagonist’, ‘LPS + TLR4 antagonist’), a random intercept per subject, and heterogeneous residual variances across groups. Residuals on the raw scale deviated clearly from normality and were therefore transformed to the log10(Pain + 1) scale. This yielded residuals that were approximately normal and homoscedastic, without systematic patterns. Estimated means and contrasts were obtained on the log scale and subsequently back-transformed to the original pain scale.

All further statistical methodology and analysis is provided in the respective supplement.

## Supporting information

Supplemental statistical methodology and analysis

## Data Availability

All data produced in the present study are available upon reasonable request to the authors.

## Acknowledgments

The authors acknowledge excellent technical support by Markus Gold-Binder and Maria Hausharter.

## Author contributions

F.R. performed the experiments, S.P. supervised the experiments. F.R., S.H. and M.J.M.F. designed the experiments; F.R., S.H. and M.J.M.F. analysed the results; SH performed the statistical analyses in R, B.J. provided support throughout the manuscript, S.S. supervised the project. F.R. S.H. and M.J.M.F drafted the manuscript, all authors contributed to the manuscript.

