## Supplemental statistical methodology and analysis for "RAGE mediates LPS-induced inflammatory pain in human skin"

### Supplementary statistical methodology and analysis

#### Transformation of injection-induced pain ratings

In accordance with the statistical analysis plan, several plausible within-subject covariance structures were evaluated, namely a compound symmetry structure (constant within-subject correlation), a first-order autoregressive structure, and low-order autoregressive moving-average structures approximating a Toeplitz covariance (the latter is not directly available in the nlme package). For each covariance structure, models with either homogeneous residual variance or injection type–specific residual variances (one variance parameter per injection type) were fitted, yielding a predefined matrix of candidate models.

Given the marked right-skewness and variance heterogeneity of the raw AUC Pain values, three transformation families were examined: $\log_{10}$(AUC Pain + 1), square-root(AUC Pain), and cubic-root(AUC Pain). Within each transformation family, all covariance structures and variance specifications were combined with the same fixed-effects structure. Candidate models were compared using the Akaike information criterion. Importantly, statistical significance (p-values, confidence intervals) was not considered at this stage; model selection was based solely on goodness-of-fit criteria and diagnostic performance. Across transformations, models based on $\log_{10}$(AUC Pain + 1) provided the most favourable balance of residual normality, reduced heteroscedasticity and interpretability on the original scale. Within the log-transformed set, the model combining a compound symmetry correlation structure with injection type–specific residual variances yielded the lowest Akaike information criterion and exhibited no systematic residual patterns. This model was therefore selected as the final working model. Estimated means for each injection type were obtained on the $\log_{10}$ scale and subsequently back-transformed to the original AUC Pain scale using the delta method–based regridding implemented in the emmeans package, yielding geometric means of AUC Pain with 95% Wald confidence intervals.

Model diagnostics were assessed for all models by inspecting the standard non-linear mixed effects diagnostic plots (residuals vs. fitted values, Q–Q plots, and leverage diagnostics). Residuals showed no systematic patterns, and variance modelling using group-specific residual variances adequately accounted for heteroscedasticity.

Modelled results of central tendencies are reported throughout the manuscript as ‘mean estimates’, despite recognizing that this measure can also be labelled as ‘estimated marginal means’, or as ‘least squares means’. Further, estimated means from mixed-effects models on log-transformed data should be interpreted as geometrically scaled after back-transformation. For readability, this is not termed ‘geometric mean estimates’ but still ‘mean estimates’.

**Model-based parametric simulation to estimate the relative effect of TRPV1 inhibition on acid-induced pain**

To determine the relative position of BCTC-containing acidic injections relative to acidic injections and pH 7.4 injections, we applied a parametric, model-based simulation that generates model-implied datasets from the fitted mixed model. This approach incorporates all estimated components of the model including fixed effects, random intercepts, condition-specific residual variances, and the within-subject compound-symmetry correlation structure to obtain a model-consistent sampling distribution of predicted pain responses for each injection type.

For this purpose, 100,000 complete datasets were simulated under the fitted model, holding all parameters at their estimated values while incorporating the model-estimated random and residual variability. In each simulation, the expected log-transformed AUC Pain for every observation was obtained from the fixed-effect component. The subject-specific random intercept was added to preserve the between-subject variability encoded in the model, and residual variation was introduced using condition-specific standard deviations. To reproduce within-subject dependence, the residuals were drawn jointly from a multivariate normal distribution parameterized to match the compound-symmetry structure estimated in the original analysis. In this way, residual noise for the repeated observations of each subject was not generated independently, but in a way that preserved the correlation pattern observed within individuals. Each simulated dataset thus represents a plausible repetition of the experiment based on the fitted mixed model. Within every simulated dataset, pain responses were summarised as geometric means for each injection type, consistent with the log-transformed modelling framework. To express where the injection type lies between two reference injection types, these simulated geometric means were mapped onto a linear 0–100% scale by assigning the lower reference (pH 7.4 injections in control or LPS) a value of 0% and the upper reference (acidic injections in control or LPS) a value of 100%. The BCTC-containing acidic injection was then positioned proportionally between these anchors (in control and LPS, respectively). Repeating this transformation across all 100,000 simulations yielded an empirical distribution of percentages relative to the references. From these simulation-derived distributions, 2.5% and 97.5% quantiles were extracted as simulation-based 95% confidence intervals.

**Modelling the time course of pain using generalized additive mixed models**

To estimate injection-type-specific differences in pain intensity continuously over time and to relate these dynamic effects to the concurrently changing pH, a generalised additive mixed model (GAM) was fitted to the raw pain ratings over time. For modelling, pain ratings were analysed on the log-transformed scale $\log_{10}$(Pain + 1) to stabilise variance and accommodate the right-skewed distribution of the raw values, while retaining interpretability on the original scale via back-transformation.

The GAM was specified to allow flexible, injection-type–specific time courses while accounting for between-subject heterogeneity and period effects. Time in seconds after injection onset was used as a continuous predictor, and a separate smooth function of time was estimated for each injection type via a factor*smooth interaction. This was implemented by estimating a separate smooth function of time for each injection type. In mgcv, this is achieved using so-called "by-smooths": each smooth term contributes only for observations belonging to its respective group and is zero otherwise. Given that the by-variable was defined as time itself for the respective injection type and zero otherwise, the contribution of each smooth is forced to be zero at time 0, such that all injection-specific curves are naturally anchored at the pre-defined fixed baseline of pain = 0 at time = 0. Each smooth was given a moderate basis dimension (k = 7), which limits the maximum number of effective bends it can exhibit and thus allows the main shape of the trajectories to be captured without overfitting noise. The effective wiggliness is then determined by penalization, so that the estimated curve uses only as much flexibility as the data support. The adequacy of k = 7 was verified using mgcv's k-index diagnostic, which showed no evidence that a larger basis (and thus a higher maximum permissible wiggliness of the smooth) would be required.

To represent subject-specific deviations from the population mean curve, a random-effect term was included as a smooth of subject identifier with a numeric-by time structure, which behaves like a random slope over time for each subject. Each subject was allowed to have their own linear trend (a deviation in slope), while no subject-specific intercept or higher-order wiggles are estimated because the by-variable constrains the smooth to be zero at time 0. This model incorporates individual differences in rate of change over time, but not full subject-specific smooth curves.

In addition, a factor*smooth interaction between time and crossover period was included as a nuisance term to adjust for potential systematic differences in pain dynamics across periods. This means that for each crossover period, the model allowed the time course to assume a slightly different shape, ensuring that any period-specific variation in pain dynamics was not inadvertently attributed to the injection types. The model was fitted using restricted maximum likelihood as implemented in the mgcv package in R.

For inference on the population-level time courses, injection type–specific least-squares mean curves (population smooths) were derived on a dense time grid covering 0–110 s in 0.5 s steps. These curves were obtained from the fitted GAM using its linear predictor matrix (the "lpmatrix" in mgcv), that is, the design matrix that maps the model coefficients to fitted values at the chosen grid points. In this step, subject-specific random effects were set to zero so that the resulting curves represent averages over subjects, and predictions were averaged over all period levels at the design-matrix level to obtain period-adjusted trajectories. Pointwise standard errors and 95% confidence intervals were then computed using the full covariance matrix of the GAM coefficients, ensuring that the uncertainty in each curve reflects the joint uncertainty in all model parameters. Finally, these predicted $\log_{10}(\text{Pain} + 1)$ values and their confidence limits were back-transformed to the original 0–100 scale, yielding injection-type–specific, period-adjusted LS-mean pain trajectories with 95% confidence bands over time.

To express pain as a direct function of the injected pH level, the population time courses were subsequently projected onto a pH-scaled x-axis. For this purpose, a smooth calibration function relating time to injected pH from the pooled pH measurements obtained during acidic injection protocols was constructed (monotone Hyman spline for the range 5-110 s). Each time point on the prediction grid was then translated into a pH value using this calibration, which allows LS-mean pain trajectories and confidence bands as a function of pH instead of time. This yielded injection type–specific LS-mean curves of pain versus pH with corresponding 95% confidence intervals on the original pain scale.

Model adequacy was evaluated by inspecting standard GAM diagnostics. Plots of standardised residuals versus fitted values and quantile–quantile plots of deviance residuals on the log-transformed scale did not reveal major deviations from model assumptions or residual patterns suggesting lack of fit.

Finally, to quantify differences in pain dynamics between selected injection types and to assess contrasts across the pH range, we derived pairwise and interaction contrasts directly from the fitted GAM. For each contrast of interest appropriate linear combinations of the GAM coefficients were constructed on a common pH grid using the linear predictor matrix. Differences in estimated means of pain between conditions, as well as their interaction contrasts, were then expressed on the original pain scale using a multivariate delta-method approximation to propagate the coefficient uncertainty through the back-transformation from $\log_{10}(\text{Pain} + 1)$.

**Posterior sampling of pH thresholds for prespecified pain differences**

To identify the pH level at which two injection types differ by a given amount of pain, a threshold-based analysis was carried out on posterior draws from the GAM. Specifically, this determined the pH value at which the model-implied difference in expected pain between two injection types first reached a threshold (e.g. 1 pain unit), together with corresponding uncertainty intervals.

Based on the fitted GAM for $\log_{10}(\text{Pain} + 1)$, first the joint sampling distribution of all model coefficients was approximated by a multivariate normal distribution with mean equal to the restricted maximum likelihood estimates and a covariance given by the unconditional coefficient covariance matrix returned by mgcv. From this distribution, 10,000 coefficient vectors were drawn, each representing one plausible realisation of the underlying population-level model. For each draw, and for each injection type, period-averaged population predictions of pain over a dense time (0.5 s) grid in the range 0−110 s were constructed. This yielded, for every posterior draw, a set of smooth, noise-free time courses of expected pain for all injection types.

For contrasts of interest ('Acidic' vs. 'Control', 'Acid' vs. 'Acid + TRPV1 antagonist', 'LPS + Acidic' vs. 'LPS', 'LPS + Acid' vs. 'LPS + Acid + TRPV1 antagonist') within each draw, the difference in expected pain between the two injection types was calculated as a function of time. For each posterior draw and each pain-difference threshold, the earliest time at which the modelled difference reached the threshold was identified by scanning through the predicted time points. If the threshold lay between two adjacent time points, the exact crossing time was obtained by linear interpolation between those neighbouring points. For each threshold and each contrast, the collection of 'first-exceedance times' across the 10,000 draws forms an empirical sampling distribution for the time at which the model-implied pain difference first attains the specified magnitude. From these distributions, the median as a central estimate and the 2.5th and 97.5th percentiles as 95% simulation-based confidence intervals were extracted. To avoid reporting thresholds for which the corresponding posterior differences were not consistently reached, we restricted the analysis to those pain-difference values that were exceeded by all posterior draws within the observed time window. For higher thresholds, some posterior difference curves never reached the required magnitude (yielding undefined first exceedance times), which precludes the computation of valid quantiles and would imply extrapolation beyond the supported data region.

To express these thresholds on the pH scale, the first exceedance times were mapped to pH using the previously constructed calibration function. Applying this calibration to the threshold crossing times yielded, for each contrast and each pain threshold, a posterior sample of pH values at which the model-implied pain difference first reached the specified magnitude. Again, median values and 95% simulation-based confidence intervals were reported, providing pH-resolved estimates and uncertainty bands for the shift in pH value generating a difference of 1-3 pain units between injection types. The difference in pH thresholds between inflamed and non-inflamed skin quantified the shift in the pH–pain relationship by inflammation.

**Analysis of pain induced by cannula insertion**

Pain ratings elicited by cannula insertion were first averaged within the four groups ('Control', 'LPS', 'LPS + RAGE antagonist', 'LPS + TLR4 antagonist'), yielding one value per group for each subject. These means were analysed with a linear mixed-effects model on the $\log_{10}$(Pain + 1) scale, with group as a fixed factor, a random intercept for subjects, and a variance-by-condition structure to allow for heteroscedastic residuals. The model was fitted by restricted maximum likelihood using the nlme package in R.

Estimated means on the $\log_{10}$ scale were obtained using emmeans, back-transformed to the original 0–100 scale, and used to form pre-specified contrasts (‘Control’ vs. ‘LPS’, ‘LPS’ vs. ‘LPS + RAGE antagonist’, ‘LPS’ vs. ‘LPS + TLR4 antagonist’). The antagonist effects were normalized onto a scale with 'Control’ as the 0% reference and ‘LPS’ as the 100% reference. For the groups containing the RAGE and the TLR4 antagonist, the fraction of the LPS-induced increase in insertion pain was calculated. This was done by comparing how much higher the back-transformed LS-mean was than in the ‘Control’ group, relative to the corresponding difference between the ‘LPS’ and ‘Control’ groups. The four estimated means are correlated and this percentage transformation is nonlinear, therefore confidence intervals for these percentages were obtained by parametric resampling from the multivariate normal distribution of the fixed effects, recomputing estimated means and percentages for each draw. The resulting empirical distributions were summarised by their medians and 95% simulation-based confidence intervals, which reflect the joint parameter uncertainty.

**Analysis of sex-dependent effects**

For all pre-specified analyses, a subanalysis was performed to investigate potential differences between female and male subjects by including ‘sex’ and its interaction with the respective factor of interest in the model.
